# Measurement of retrieved chunk quality from real-world knowledge in retrieval-augmented generation: A Phase 1 foundational study

**DOI:** 10.64898/2026.01.01.26343326

**Authors:** Yasuko Fukataki, Wakako Hayashi, Megumi Kitayama, Yoichi M. Ito

## Abstract

Retrieval-augmented generation (RAG) holds promise for supporting high-stakes medical decision-making. However, most research has focused on downstream optimization of parameters and algorithms. This Phase 1 foundational study quantitatively evaluated the upstream quality of knowledge documents and their impact on retrieval performance, using Japanese clinical research protocol manuals for Institutional Review Board pre-screening support as a case study. We established a three-tier evaluation framework: Level 1 assessed knowledge document quality through independent expert review across Structure, Granularity, and Noise dimensions; Level 2a evaluated the structural quality of retrieved chunks using large language model-as-a-Judge across five metrics; and Level 2b conducted proof-of-concept content appropriateness evaluation against a Gold Standard derived from international guidelines. Using Google Cloud Vertex AI Search, we analyzed 594 chunks from baseline knowledge (A-line: four institutional manuals as-is) and six chunks from optimized knowledge (B-line: proof of concept). Level 2a evaluations employed deterministic settings with five independent trials, achieving excellent reliability (intraclass correlation coefficient of 0.936). The results revealed substantial quality limitations in the A-line chunks: the median scores were 2.0 or below across all five metrics, with fewer than 20% of the chunks reaching practical utility thresholds (score of 4 or higher). Even among the top-ranked results, fewer than half met the practical utility criteria, except for Faithfulness. The inter-rater agreement in the Level 1 evaluation was fair (Fleiss kappa value of 0.269), indicating the need for framework refinement. The retrieved chunk lengths significantly exceeded the configured settings (median of 3,861 characters versus 500 tokens), potentially indicating information dilution. The B-line optimization achieved perfect scores across all metrics, demonstrating potential for improvement. These findings demonstrate that upstream document quality constrains retrieval performance, challenging assumptions regarding plug-and-play RAG deployment.

**Author Summary:** Artificial intelligence systems that retrieve information from documents and generate responses are increasingly being used to support medical decision-making. We questioned the assumption that uploading existing documents is sufficient for fine-tuning the algorithms of these systems by investigating whether document quality is a limiting factor. We studied Japanese clinical research manuals used for research ethics review, assessing the efficacy of an AI system in retrieving information from these documents. We evaluated nearly 600 text segments retrieved by the system and found that fewer than one in five segments met our quality standards, even among the highest-ranked results. The system frequently retrieved excessively long passages obscuring key information. However, when we restructured one document section using clearer organization and formatting, the system achieved perfect performance scores. This improvement suggests that not only algorithm optimization but also document preparation is crucial for system effectiveness. Our findings challenge the “plug-and-play” assumption commonly used in AI deployment. For high-stakes medical applications, organizations cannot simply expect reliable results to be obtained by uploading existing documents. Instead, they must invest in preparing well-structured knowledge documents. This foundational work establishes measurement methods to guide such preparation, which is essential before these systems can safely support healthcare decision-making.

## 1. Introduction

The rapid development of large language models (LLMs) has heightened expectations for LLM-based support systems in the medical field. However, LLMs face challenges such as hallucinations, which generate information not grounded in facts; obsolescence, wherein they lack knowledge beyond the temporal scope of their training data; and lack of specialized knowledge in specific domains [1]. Retrieval-augmented generation (RAG) has attracted attention as an approach to address these challenges. RAG is a framework that retrieves related information from external knowledge bases and utilizes the results as context for generation [2]. In particular, in the high-stakes medical field, where decision-making errors lead to serious consequences, the tolerance for hallucinations is extremely low, and high safety standards and strict control are required [3–5]. Utilizing organization-specific guidelines and manuals as RAG knowledge is expected to enable more accurate and context-appropriate support.

Research aimed at improving RAG performance has focused primarily on chunk size optimization [6], document segmentation [7, 8], and optimization of the entire RAG pipeline, including search algorithms and prompt design [9]. In recent years, cloud-based RAG platforms have been equipped with functions that automate processes ranging from layout analysis of PDFs and other documents to chunk division and indexing. The spread of such automatic ingestion functions has reinforced the practical assumption that sufficient search accuracy and practical answers can be obtained simply by inputting existing manuals and regulatory documents as they are. Conversely, some studies have highlighted the importance of upstream factors, such as document structure and knowledge base preparation [10–12], and there is growing awareness that RAG performance depends on the quality of structuring and preprocessing of input knowledge. However, despite these contrasting perspectives—specifically, the practical assumption that existing documents can be used as-is to achieve satisfactory results versus the research finding that knowledge quality strongly influences performance—the quantitative evaluation of the quality of input documents (upstream quality) at the chunk level has not been sufficiently investigated. In particular, for highly specialized and complex medical-related documents, the validity of the assumption that simply inputting existing documents yields sufficient performance has not been systematically verified yet.

The standardization of institutional review board (IRB) review processes is an internationally important issue, and multiple studies have highlighted the variability in the consistency of review criteria and evaluation [13–15]. In the context of clinical research in Japan, the review of research ethics by IRBs and certified review boards (CRBs) is mandated based on Good Clinical Practice (GCP) and the Clinical Research Act. However, materials from the Ministry of Health, Labour and Welfare’s council repeatedly report significant variability in the level of detail, structure, and evaluation perspectives of CRB review comments [16–21]. This non-uniformity is believed to be caused by the ambiguity of review criteria and the subjectivity of judgments, and standardization is required to ensure the consistency and transparency of the review process.

IRB pre-screening requires not only compliance with formal requirements of study protocols (presence or absence of descriptions of required items) but also a substantive evaluation of the scientific and ethical validity of the described content. Even in the future construction of a support system using RAG technology, an important point of discussion is how to assist in judging the validity of protocol content in addition to meeting formal requirements.

However, the preconditions for implementing such advanced validity-assessment RAG have not been sufficiently established. Specifically, three points are lacking: (1) standardized evaluation criteria, (2) formalization of tacit knowledge of review committee members, and (3) LLM-interpretable structured knowledge. In particular, the criteria for evaluating knowledge quality depend on subjective judgments that differ in interpretation, even among experts, and a framework to ensure the consistency of evaluation has not been established. Study protocol development manuals independently established by each medical institution exhibit non-uniform structure and description granularity. Consequently, utilizing these manuals as they are as RAG knowledge does not necessarily enable a consistent validity assessment. Therefore, the development of a validity-assessment RAG requires a foundational study to quantify the extent to which current manuals qualify as RAG knowledge, verify the validity of the evaluation criteria, and clarify issues associated with the formalization of tacit knowledge and the development of an LLM-interpretable knowledge base infrastructure. This work is the first foundational study (Phase 1) of a planned multi-phase project for implementing a validity-assessment RAG in the future.

As a first step toward the future development of a validity-assessment RAG for protocol content, this study aimed to quantitatively evaluate the actual state of upstream knowledge quality at the retrieval stage of RAG, specifically targeting Japanese study protocol development manuals, and identify its structural issues and potential areas for improvement. In this Phase 1 foundational study, we first (1) quantified the quality distribution of raw chunks (A-line) retrieved under the standard settings of Google Cloud Vertex AI Search. Subsequently, (2) we constructed a three-tier evaluation framework consisting of expert evaluation (Level 1), LLM-as-a-Judge (Level 2a), and Gold Standard (GS)-based evaluation (Level 2b). Furthermore, (3) by comparing the A-line (knowledge inputted as-is from existing manuals) and B-line (structured knowledge), we exploratorily examined the direction of upstream quality improvement.

As a Phase 1 foundational study, this work established a systematic framework for evaluating upstream knowledge quality at the retrieval stage of RAG systems under realistic, unrefined knowledge conditions, and clarified its feasibility and remaining challenges.

In this study, we designed a three-tier evaluation framework, examined its validity by identifying issues in the evaluation criteria through the measurement of inter-rater agreement, explored the reliability of LLM-as-a-Judge, and presented quantitative suggestions showing the potential for improvement through structured knowledge. These results contribute to the design of the next phase, which aims to formalize the tacit knowledge of the review committee members and develop an LLM-interpretable knowledge base infrastructure.

## 2. Results

In this study, to comprehensively evaluate the upstream quality of RAG systems, we implemented a three-tier evaluation framework comprising Level 1: knowledge quality evaluation by domain experts, Level 2a: structural quality evaluation of chunks, and Level 2b: content appropriateness evaluation of chunks. We extracted 594 chunks from the A-line and six chunks from the B-line. Following test–retest reliability verification (intraclass correlation coefficient, ICC) by LLM-as-a-Judge, we implemented descriptive statistics, Top-K monotonicity analysis, knowledge quality evaluation, and two proofs of concept (B-line structural optimization and L2b GS evaluation). The Level 2b evaluation targeted 99 chunks of the Include query type and QE=true condition extracted from the A-line. In this section, we report the measurement properties and distribution of quality metrics.

### 2.1 Overview of retrieved chunks

The overall composition of the chunks targeted for evaluation in this study is presented. The theoretical total number of chunks in the experimental design was 720 (four sites × nine queries × two QE conditions × 10 ranks). However, in some knowledge documents, the amount of text in specific sections was less, resulting in cases where the number of chunks returned as search results per query × QE condition did not reach the upper limit of 10. Owing to this application programming interface (API) specification constraint, the number of chunks actually extracted and analyzed in the A-line was 594.

These chunks comprised combinations of three sections (Objective, Design, Risks_benefits) × four sites (site-01 to site-04) × two query expansion conditions (QE on/off) × maximum 10 ranks. Conversely, in the B-line, only the Objective section was targeted, and six chunks were extracted from combinations of three query types × two query expansion conditions.

In the Level 2a evaluation (structural quality evaluation), all 600 chunks (594 A-line and six B-line chunks) were targeted. In the Level 2b evaluation (content appropriateness evaluation), 99 chunks (Objective: 33, Design: 33, Risks_benefits: 33) were extracted by stepwise filtering (line=“A”, query_type=“include”, qe=true).

### 2.2 Reliability of LLM-as-a-Judge for structural quality assessment

The test–retest reliability of the measurement instrument (LLM-as-a-Judge) in the Level 2a evaluation was evaluated using ICC. Five independent evaluations were each implemented for the 594 A-line chunks, and a total of 2,970 evaluations were targeted. Missing scores were not observed, and as a result of calculating ICC(2,1) with all 2,970 evaluations as analysis targets, the preset gate condition (ICC ≥ 0.75, Good or above) was met for all five metrics.

The overall ICC (five-metric mean score) was 0.936 (95%confidence interval (CI): 0.930– 0.940) and was judged as Excellent. The ICC by metric was as follows: Section Targeting: 0.964 (95%CI: 0.960–0.970), Coverage: 0.951 (95%CI: 0.950–0.960), Purity: 0.882 (95%CI: 0.870–0.900), Granularity: 0.755 (95%CI: 0.730–0.780), and Faithfulness: 0.859 (95%CI: 0.840–0.870). Section Targeting and Coverage were judged as Excellent, whereas Purity, Granularity, and Faithfulness were judged as Good.

The ICC of all metrics corresponded to Good or above (≥ 0.75), thereby confirming the test–retest reliability of LLM-as-a-Judge; therefore, in the subsequent main analysis, the five-trial mean score of each chunk was used. Note that the six B-line chunks were positioned as proof of concept and were excluded from ICC verification owing to sample size constraints (Table 1).

**Table 1.** ICC(2,1) of chunk structural quality scores in Level 2a.

| Metric | ICC(2,1) | CI95_lower | CI95_upper | Interpretation |
| --- | --- | --- | --- | --- |
| Overall | 0.936 | 0.93 | 0.94 | Excellent |
| section_targeting | 0.964 | 0.96 | 0.97 | Excellent |
| coverage | 0.951 | 0.95 | 0.96 | Excellent |
| purity | 0.882 | 0.87 | 0.9 | Good |
| granularity | 0.755 | 0.73 | 0.78 | Good |
| faithfulness | 0.859 | 0.84 | 0.87 | Good |

### 2.3 Structural quality of retrieved chunks (Level 2a)

#### 2.3.1 Overall quality distribution

Upon evaluating the structural quality of the 594 A-line chunks using five metrics, the medians were as follows: Section Targeting: 0.0, Coverage: 0.0, Purity: 1.0, Granularity: 2.0, Faithfulness: 0.0. The practical utility metric P(score ≥ 4) yielded the following results: Section Targeting: 11.8% (95%CI: 9.4–14.6%), Coverage: 10.8% (95%CI: 8.5–13.5%), Purity: 5.6% (95%CI: 4.0–7.7%), Granularity: 5.9% (95%CI: 4.3–8.1%), Faithfulness: 16.8% (95%CI: 14.0–20.1%).

For all five metrics, the median was 2.0 or below, and less than 20% of the chunks achieved the practical utility level (score ≥ 4). Faithfulness showed the highest P(score ≥ 4) of 16.8% but remained at a low level. The standard deviation ranged from 1.1 to 1.8, reflecting substantial variability in chunk quality (Table 2, Fig 1).

**Fig 1.**
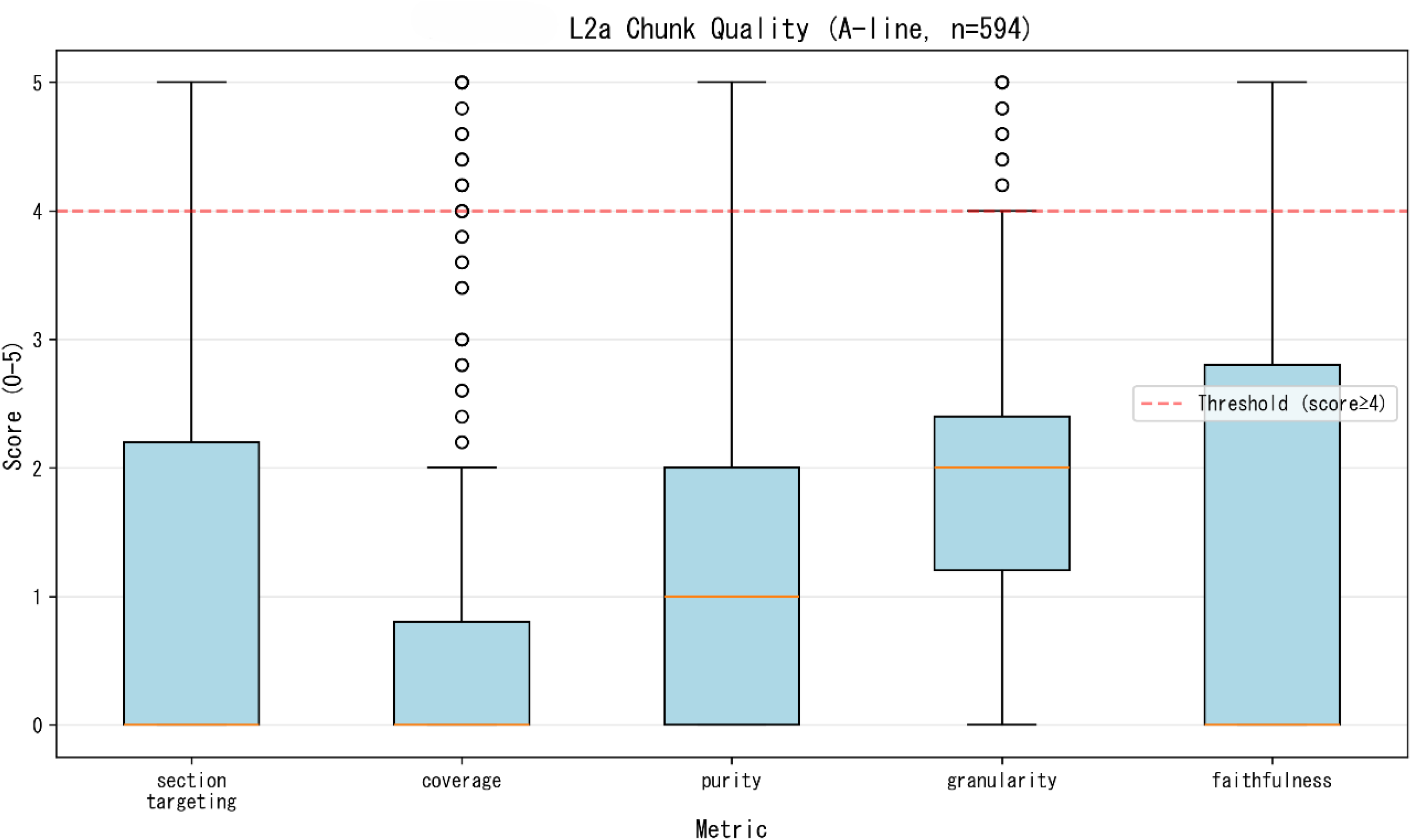
Distribution of Level 2a structural quality scores in the 594 A-line chunks Each box plot shows the score distribution (0–5) of five metrics (Section Targeting, Coverage, Purity, Granularity, and Faithfulness), and the dashed line shows the practical utility threshold (score ≥ 4).

**Table 2.**
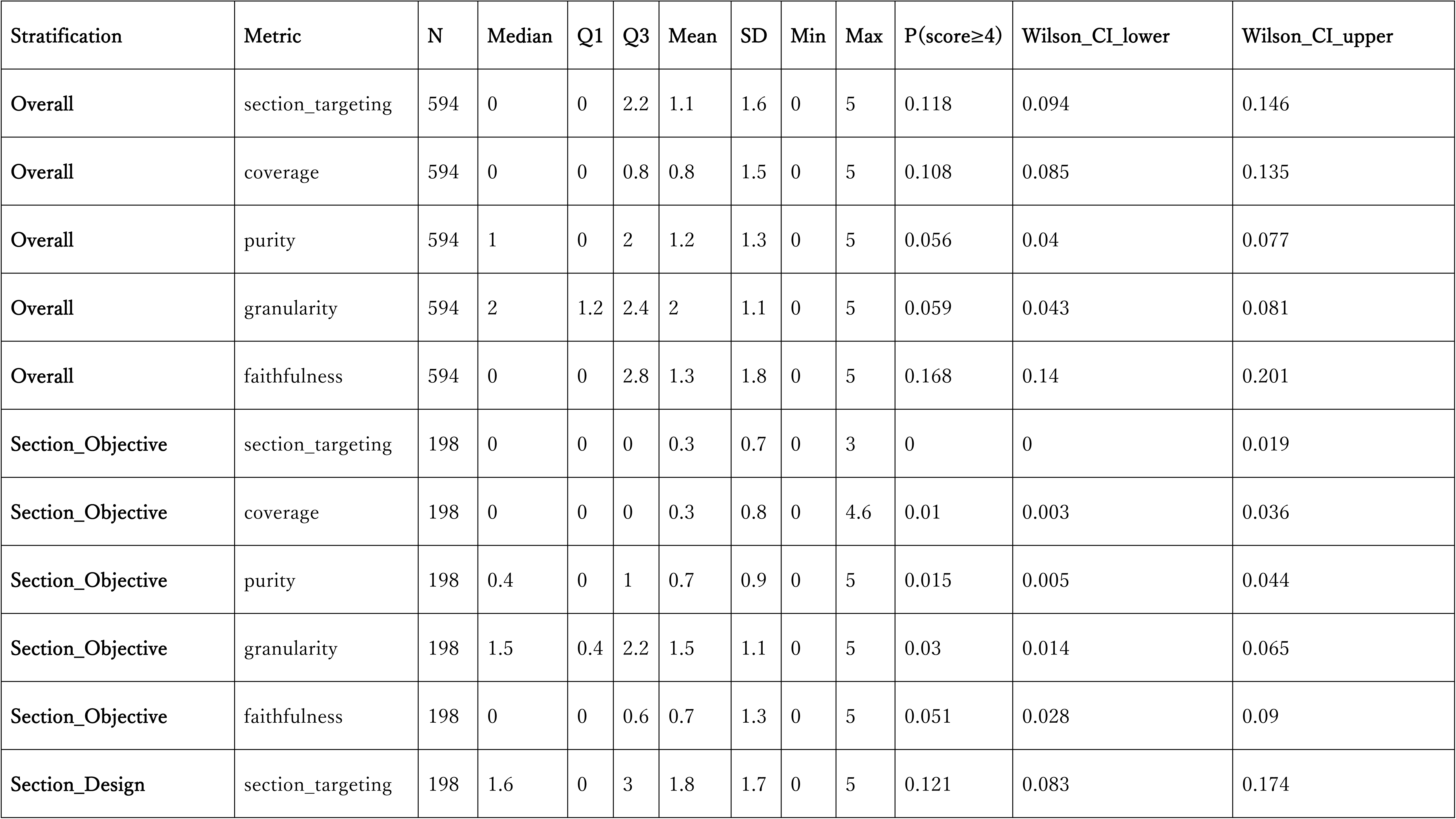
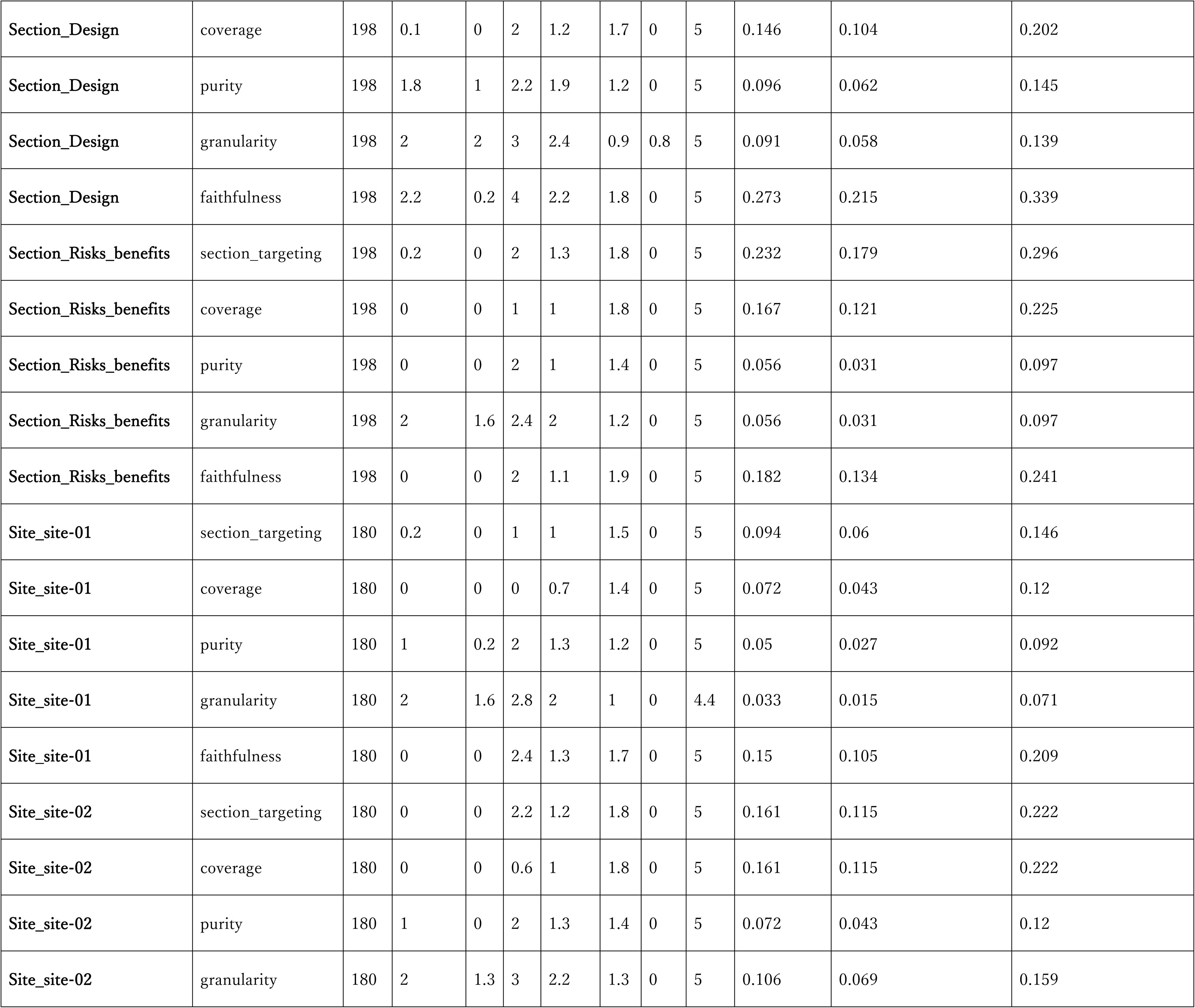
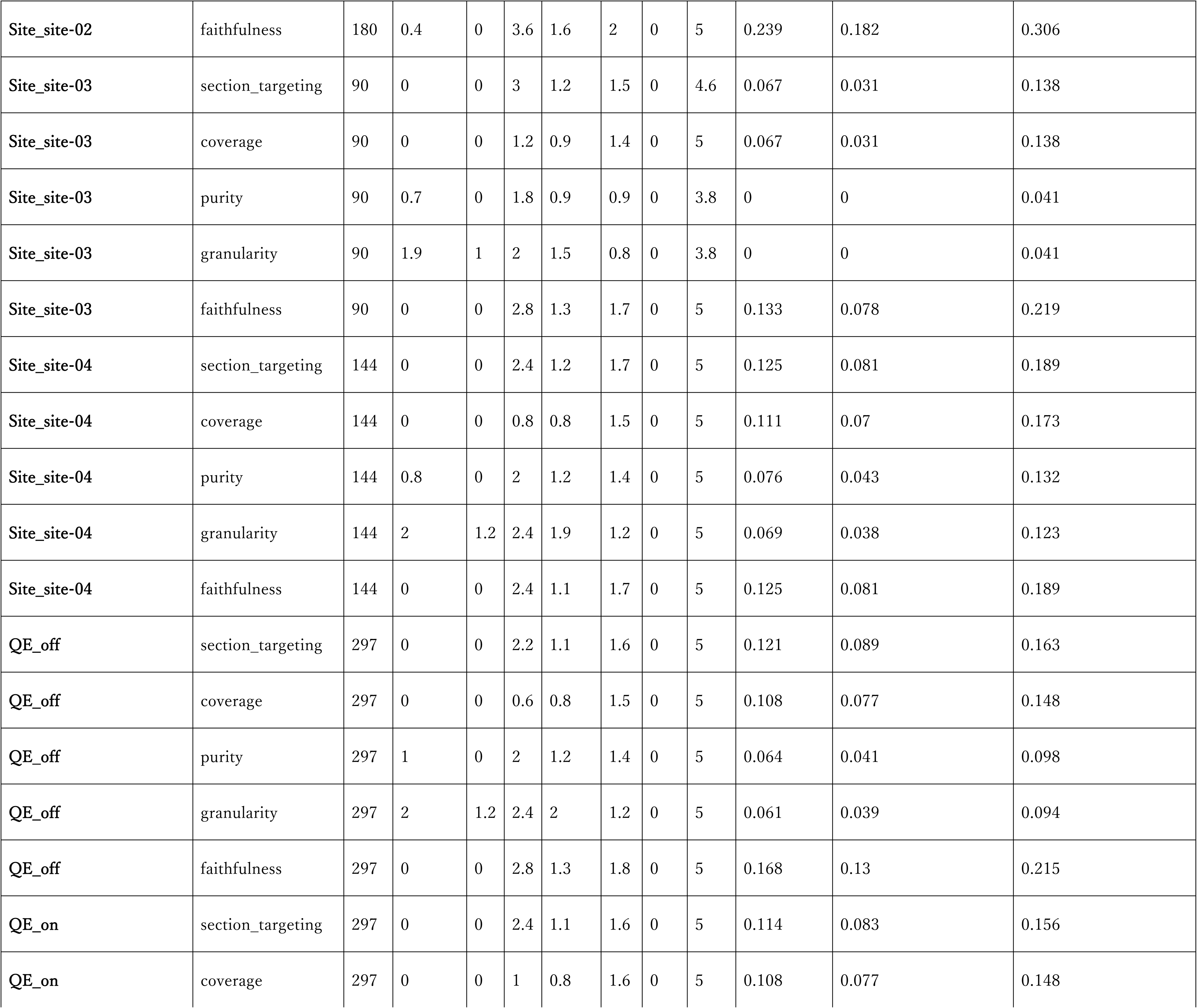
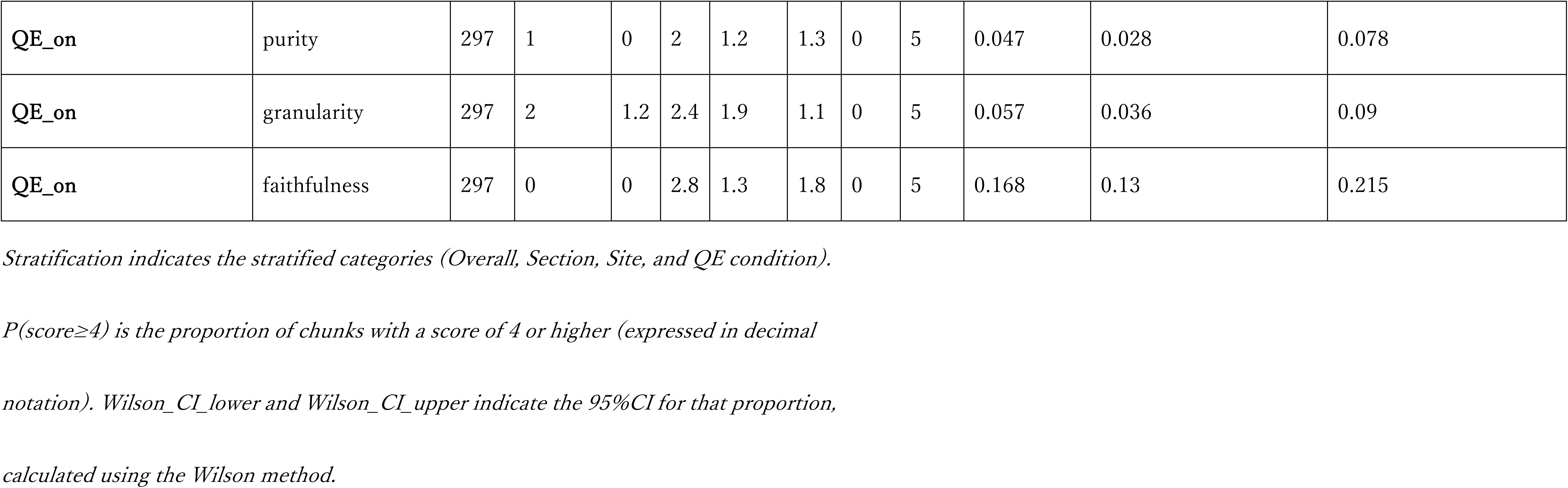
Descriptive statistics of chunk structural quality scores in Level 2a (A-line, n = 594)

#### 2.3.2 Quality variation by section

Stratified analysis by section revealed notable differences in the distribution of the quality scores (Table 2, Fig 2). Objective (n=198) was the lowest across all metrics, showing a median of 0.0 for Section Targeting and P(score ≥ 4) of 0.0%. Design (n=198) was relatively high, showing medians of 1.6 and 2.2 for Section Targeting and Faithfulness, respectively, and P(score ≥ 4) of Faithfulness reached 27.3% (95%CI: 21.5–33.9%).

**Fig 2.**
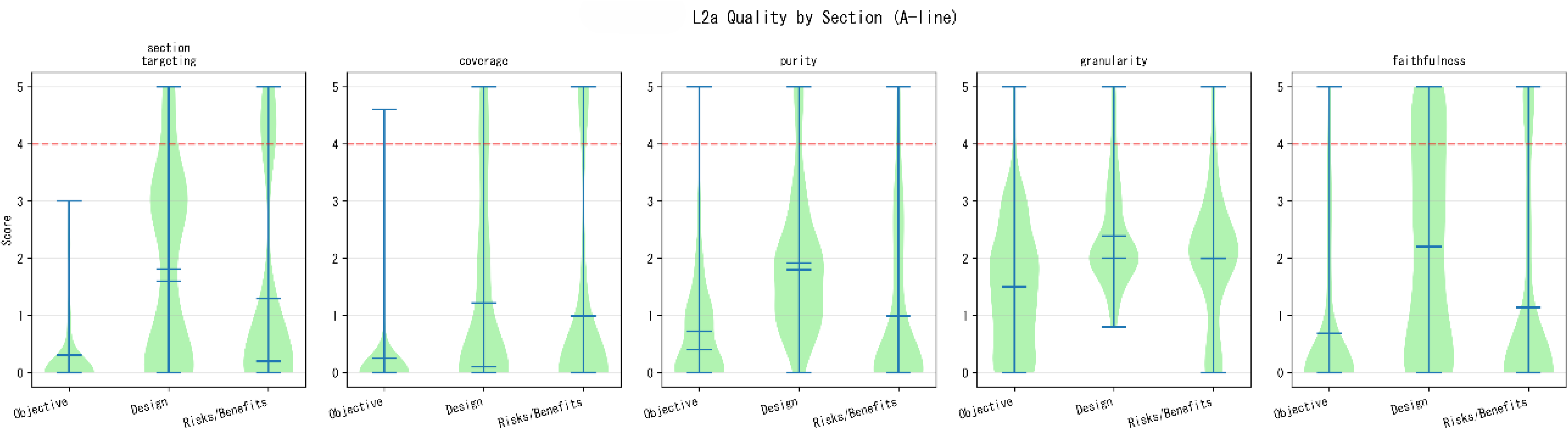
Distribution of structural quality scores by section in the A-line chunks (Level 2a) Each violin plot shows the score distribution (0–5) of five metrics (Section Targeting, Coverage, Purity, Granularity, and Faithfulness), and the horizontal axis shows three sections (Objective, Design, and Risks_benefits). The red dashed line indicates the practical utility threshold (score ≥ 4).

Risks_benefits (n=198) showed different patterns depending on the metric. P(score ≥ 4) of Section Targeting was 23.2% (95%CI: 17.9–29.6%), the highest among the three sections. However, for Coverage, the median was 0.0, and P(score ≥ 4) was 16.7% (95%CI: 12.1–22.5%), comparable to Design (14.6%, 95%CI: 10.4–20.2%). For Faithfulness, the median was 0.0, and P(score ≥ 4) was 18.2% (95%CI: 13.4–24.1%), showing intermediate values.

#### 2.3.3 Quality variation by site and query expansion

Stratified analysis by site (site-01 to site-04) and QE (on/off) was performed (Table 2). Site-02 showed the highest P(score ≥ 4) of 16.1% (95%CI: 11.5–22.2%) for Section Targeting and Coverage, and site-03 showed the lowest across all metrics, with P(score ≥ 4) of Purity and Granularity at 0.0% (95%CI: 0.0–4.1%).

QE=on (n=297) showed medians of 0.0–2.0, and QE=off (n=297) showed medians of 0.0–2.0. No notable differences were observed between the two conditions. P(score ≥ 4) of Purity was 4.7% (95%CI: 2.8–7.8%) for QE=on and 6.4% (95%CI: 4.1–9.8%) for QE=off, and no quality improvement effects were observed by enabling query expansion.

### 2.4 Chunk length distribution

The character count distribution of actually retrieved chunks was measured against the chunk size setting (chunkSize=500 tokens) of Vertex AI Search.

#### 2.4.1 Overall distribution

The character count distribution of the 594 A-line chunks exhibited a range of 62–10,343 characters and a long right tail (Fig 3). The mode was located within the 0–500 character bin, with approximately 84 chunks (14.1%) fitting near the set value. However, most chunks significantly exceeded the set value. The distribution exhibited a second peak at approximately 2,000–3,000 characters, and numerous long chunks of 6,000 characters or more were observed.

**Fig 3.**
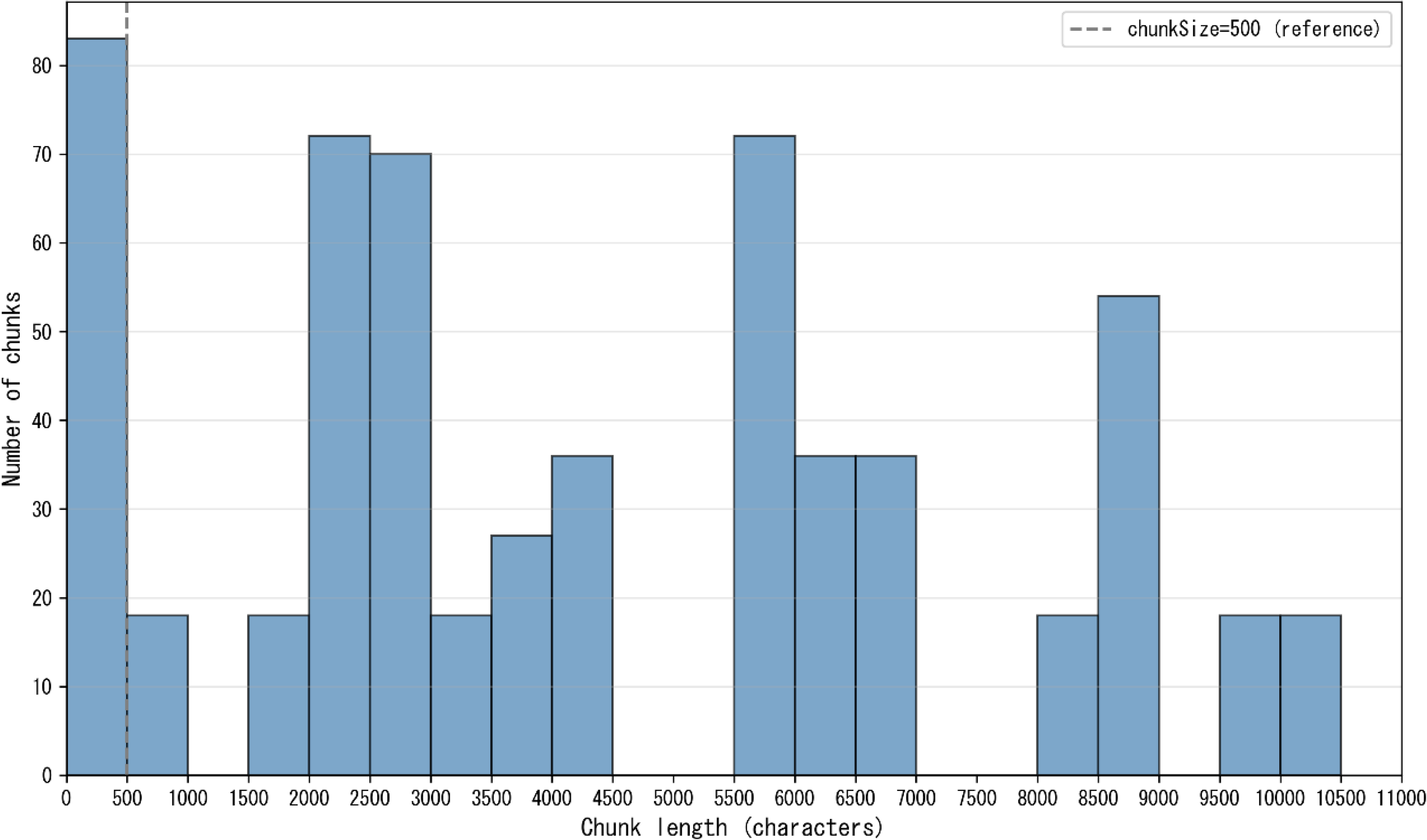
Character count distribution of the 594 A-line chunks (histogram) The horizontal axis represents the chunk character count, and the vertical axis represents the number of chunks included within each bin. The dashed line serves as a reference for the set chunkSize (equivalent to 500 tokens).

#### 2.4.2 Variation by QE and section

The chunk length distributions were stratified by six QE × Section categories (Table 3, Fig 4). Across all categories, the retrieved chunks were consistently large and substantially exceeded the configured chunkSize setting (500 tokens).

**Fig 4.**
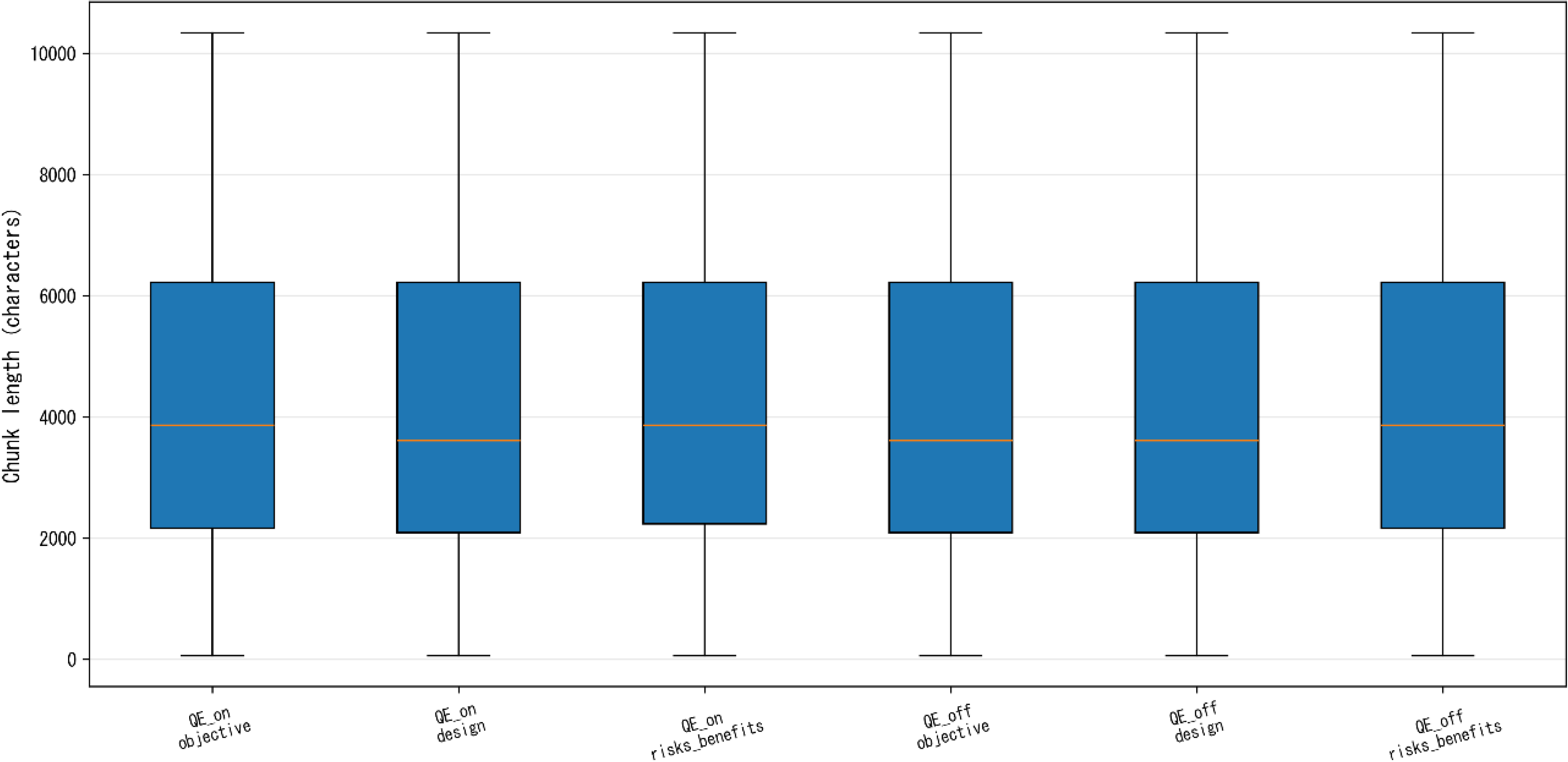
Chunk character count distribution by QE × section (box plot) The character count distribution is shown for each combination of QE_on/off and the three sections (Objective/Design/Risks_benefits). The box represents the IQR, center line represents the median, and whiskers represent the minimum and maximum values.

**Table 3.**
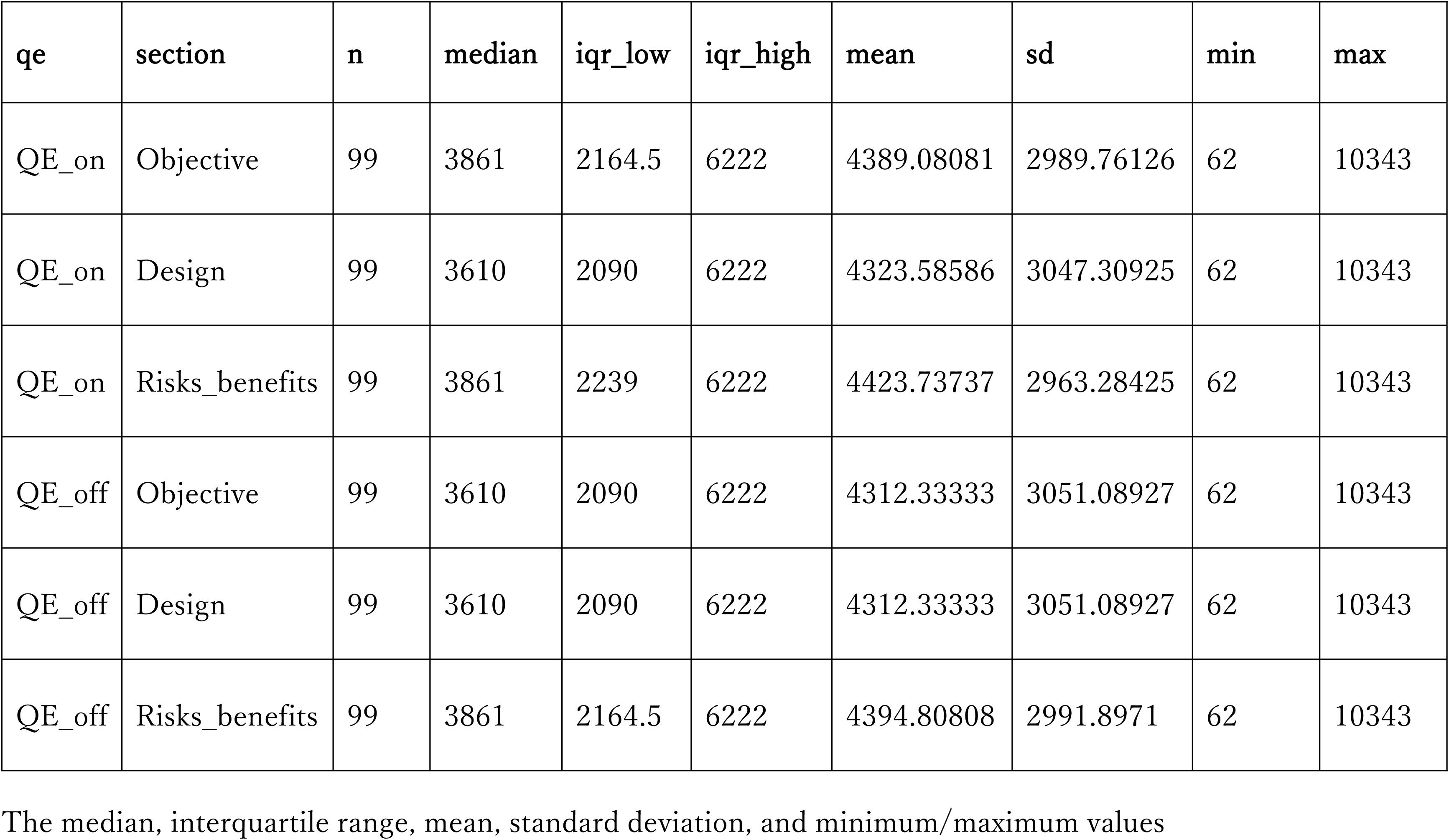
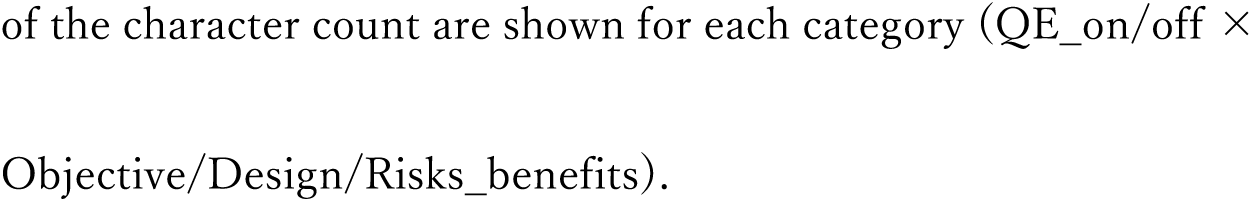
Descriptive statistics of character counts of the 594 A-line chunks (categorized by QE condition × section)

The median chunk lengths were approximately 3,600–3,900 characters across all categories, and the mean values ranged from approximately 4,300 to 4,400 characters. The upper bounds of the interquartile ranges were identical across all categories (6,222 characters), and the minimum and maximum values were also observed within the same ranges.

No marked differences in chunk length distributions were observed between QE conditions (on vs. off) or among sections (Objective, Design, and Risks_benefits).

### 2.5 Relationship between retrieval rank and chunk quality

#### 2.5.1 Main analysis: Rank 1–5 monotonicity

To verify the monotonic tendency that a smaller retrieval rank (Rank) corresponds to a higher chunk quality score, the Jonckheere–Terpstra test and Kendall’s *τ* were implemented to 360 chunks of Rank 1–5 (Rank 1 group: n=72, Rank 2–3 group: n=144, Rank 4–5 group: n=144; Table 4).

**Table 4.**
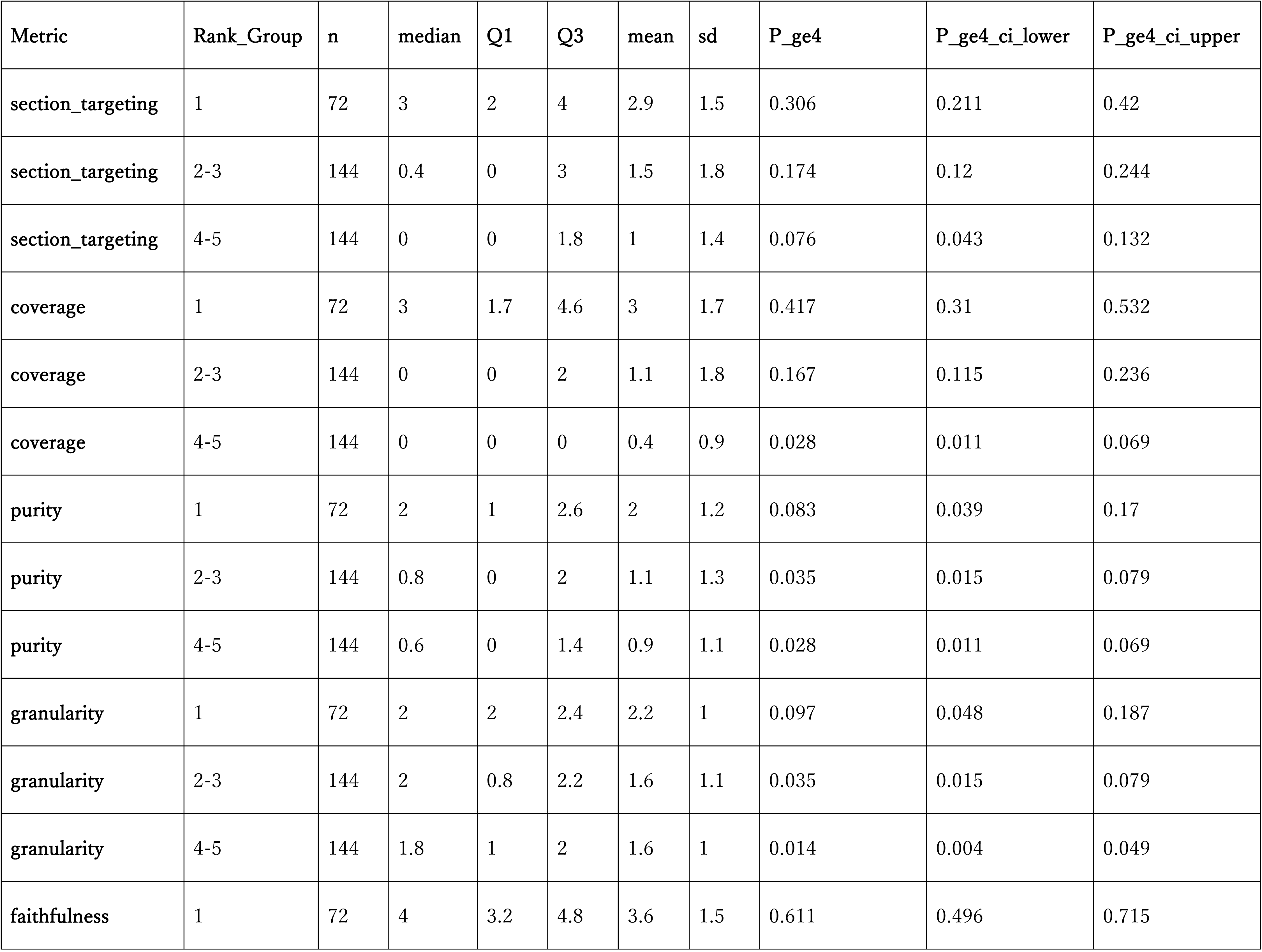
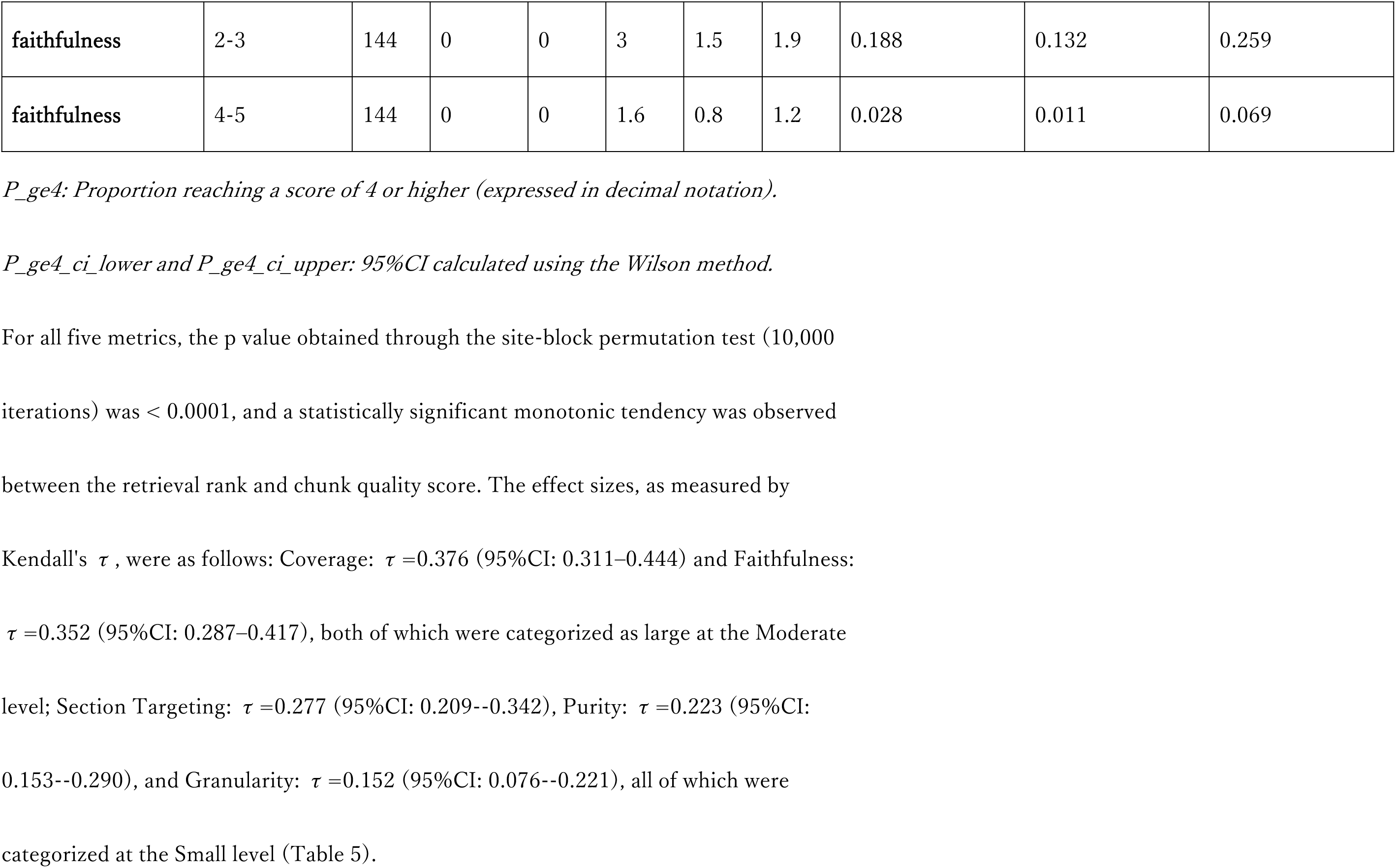
Descriptive statistics of chunk quality scores categorized by Rank group (Rank 1–5, n=360)

**Table 5.**
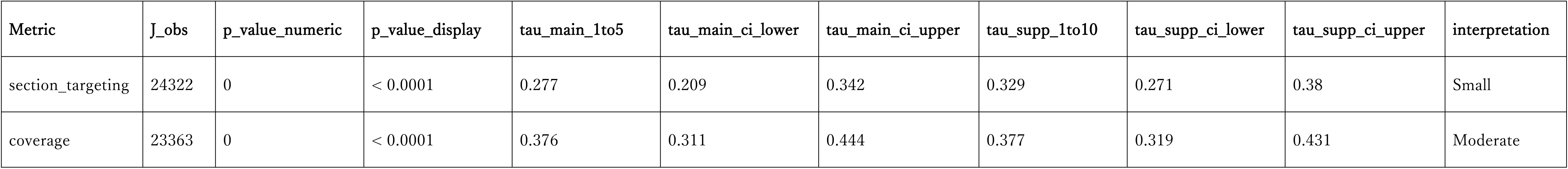
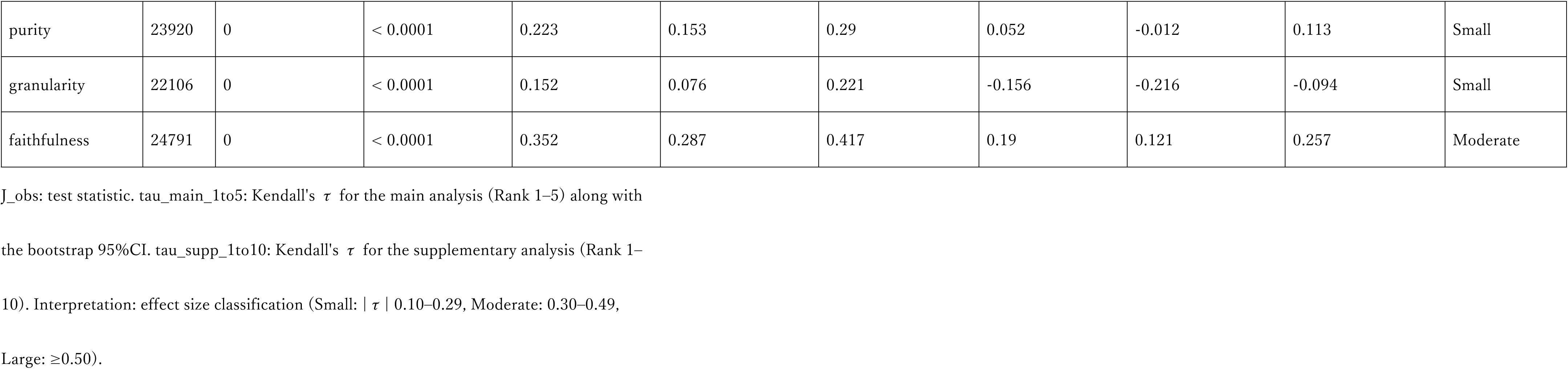
Monotonic relationship between retrieval rank and chunk quality (Jonckheere–Terpstra test)

In the group-wise descriptive statistics (Table 4, Fig 5), the Rank 1 group showed the highest median and P(score ≥ 4) across all metrics. For Faithfulness, Rank 1 group exhibited a median of 4.0 (IQR: 3.2–4.8) and P(score ≥ 4) of 61.1% (95%CI: 49.6–71.5%), whereas Rank 4–5 group exhibited a median of 0.0 (IQR: 0.0–1.6) and P(score ≥ 4) of 2.8% (95%CI: 1.1–6.9%). For Coverage, Rank 1 group exhibited a median of 3.0 (IQR: 1.7–4.6) and P(score ≥ 4) of 41.7% (95%CI: 31.0–53.2%), whereas Rank 4–5 group exhibited a median of 0.0 (IQR: 0.0–0.0) and P(score ≥ 4) of 2.8% (95%CI: 1.1–6.9%).

**Fig 5.**
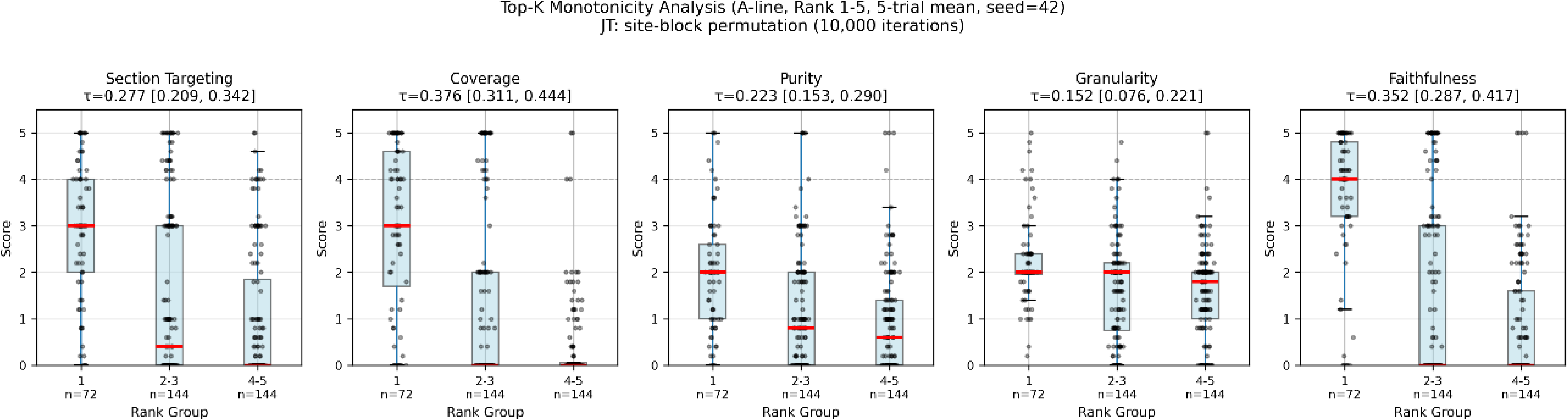
Chunk quality score distribution at Rank 1–5 (5 metrics) Total 360 A-line chunks (Rank 1–5) were divided into three groups (Rank 1, Rank 2–3, and Rank 4–5), and the score distribution across five structural quality metrics (Section Targeting, Coverage, Purity, Granularity, and Faithfulness) is illustrated using box plots. The values of *τ* and 95%CI at the top of each panel show the estimate of Kendall’s *τ* (main analysis: Rank 1–5). The box represents the IQR, center line represents the median, whiskers represent the minimum and maximum values, and dots represent individual chunks. The gray dashed line shows the predefined practical utility level threshold (score ≥ 4).

However, even within the Rank 1 group, the proportion of chunks that reached the predefined practical utility level (score ≥ 4) was limited. For the four metrics, excluding Faithfulness, P(score ≥ 4) in the Rank 1 group remained in the range of 8.3–41.7%. Notably, Purity and Granularity exhibited particularly low values of 8.3% (95%CI: 3.9– 17.0%) and 9.7% (95%CI: 4.8–18.7%), respectively (Table 4).

#### 2.5.2 Supplementary analysis: Rank 1–10 detail

To ensure transparency, a supplementary analysis was performed on all 594 A-line chunks (Rank 1–10). At Rank 6–10, the sample size decreased (n=54 or n=36; S1 Table), which requires cautious interpretation.

Kendall’s *τ* for the overall Rank 1–10 (Table 5) showed different patterns for some metrics compared to the main analysis (Rank 1–5). Coverage and Section Targeting maintained positive correlations, with *τ*=0.377 (95%CI: 0.319–0.431) and 0.329 (95%CI: 0.271–0.380), respectively, consistent with the main analysis. By contrast, for Granularity, while the main analysis exhibited a positive correlation with *τ*=0.152, Rank 1–10 exhibited a negative correlation with *τ*=−0.156 (95%CI: −0.216–−0.094). For Purity, while the main analysis yielded *τ*=0.223, Rank 1–10 exhibited *τ*=0.052 (95%CI: −0.012–0.113), with the effect size significantly decreasing and the CI including 0.

In the P(score ≥ 4) distribution by Rank (S1 Table, S1 Fig), Rank 1 exhibited the highest values across all metrics. However, at Rank 10, certain metrics showed higher values compared to Rank 2–9: Granularity: 38.9% (95%CI: 24.8–55.1%), Faithfulness: 38.9% (95%CI: 24.8–55.1%), and Purity: 25.0% (95%CI: 13.8–41.1%) (S2 Fig).

### 2.6 Knowledge quality evaluation by domain experts (Level 1)

#### 2.6.1 Inter-rater agreement (Fleiss’ kappa)

For independent evaluation by three domain experts (160 evaluation units = four sites × 40 items) involving knowledge documents from four A-line sites, inter-rater agreement was evaluated using Fleiss’ kappa (Table 6).

**Table 6.**
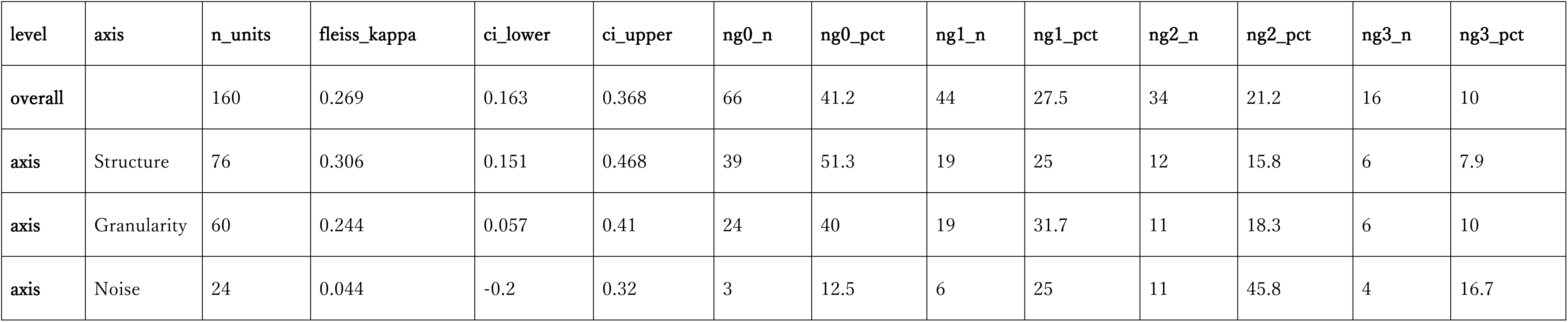
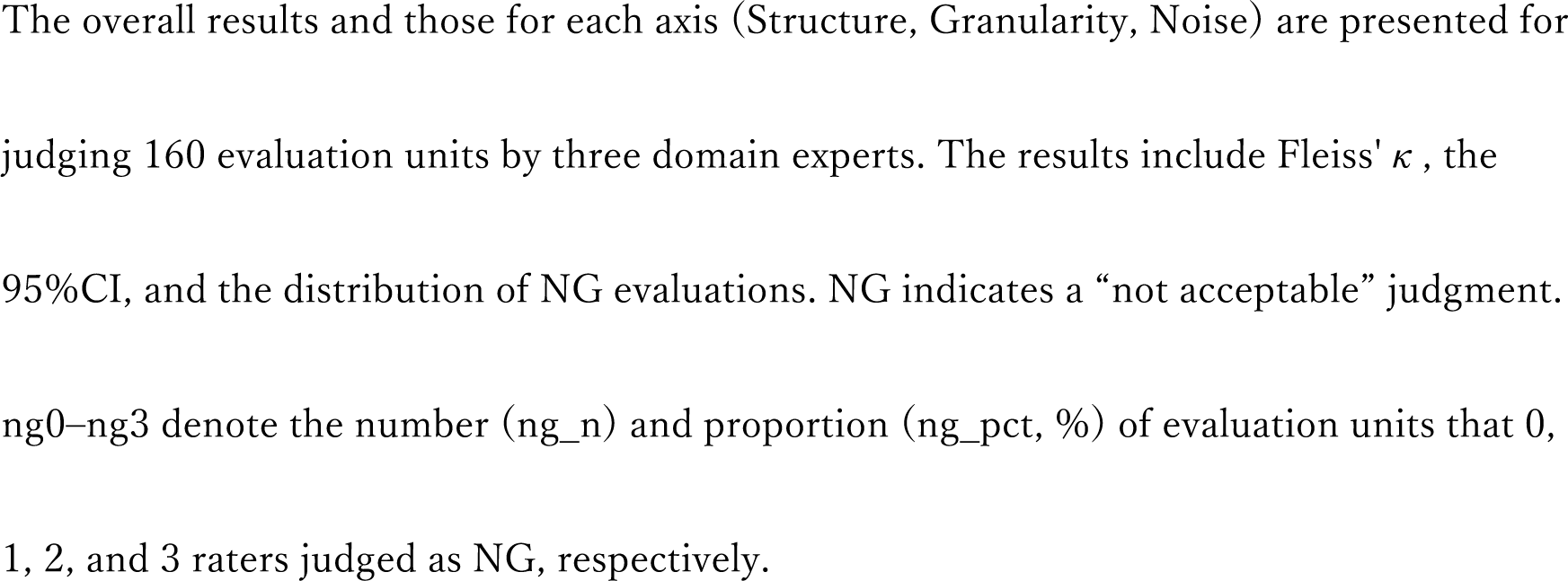
Expert evaluation for A-line knowledge quality (Level 1)

Overall, Fleiss’ kappa was 0.269 (95%CI: 0.163–0.368). The values by axis are as follows: Structure axis (n=76): *κ*=0.306 (95%CI: 0.151–0.468), Granularity axis (n=60): *κ*=0.244 (95%CI: 0.057–0.410), and Noise axis (n=24): *κ*=0.044 (95%CI: −0.200–0.320). The 95%CI of the Noise axis included 0 (95%CI: −0.200–0.320).

#### 2.6.2 Distribution of NG judgments

For each evaluation unit (site–item pair), the number of raters among the three experts who judged the unit as NG was tallied (Table 6). Overall (n=160), the results were as follows: NG×0 (all OK): 66 (41.2%), NG×1 (only one NG): 44 (27.5%), NG×2 (two NG): 34 (21.2%), and NG×3 (all NG): 16 (10.0%).

In the NG distribution by axis, the Structure axis (n=76) had NG×0 at 51.3%, which was the highest, indicating a tendency among raters to agree easily. Conversely, the Noise axis (n=24) exhibited a low NG×0 at 12.5%, whereas NG×2 represented the highest proportion at 45.8%, indicating a tendency toward judgment divergence among raters. The Granularity axis (n=60) showed an intermediate distribution with NG×0 at 40.0%, NG×1 at 31.7%, NG×2 at 18.3%, and NG×3 at 10.0%.

### 2.7 Proof-of-concept: Optimized knowledge structure (B-line)

#### 2.7.1 B-line descriptive statistics

Descriptive statistics using five-trial mean scores were calculated for six chunks (Objective section only) that were targets of the B-line evaluation (Table 7).

**Table 7.**
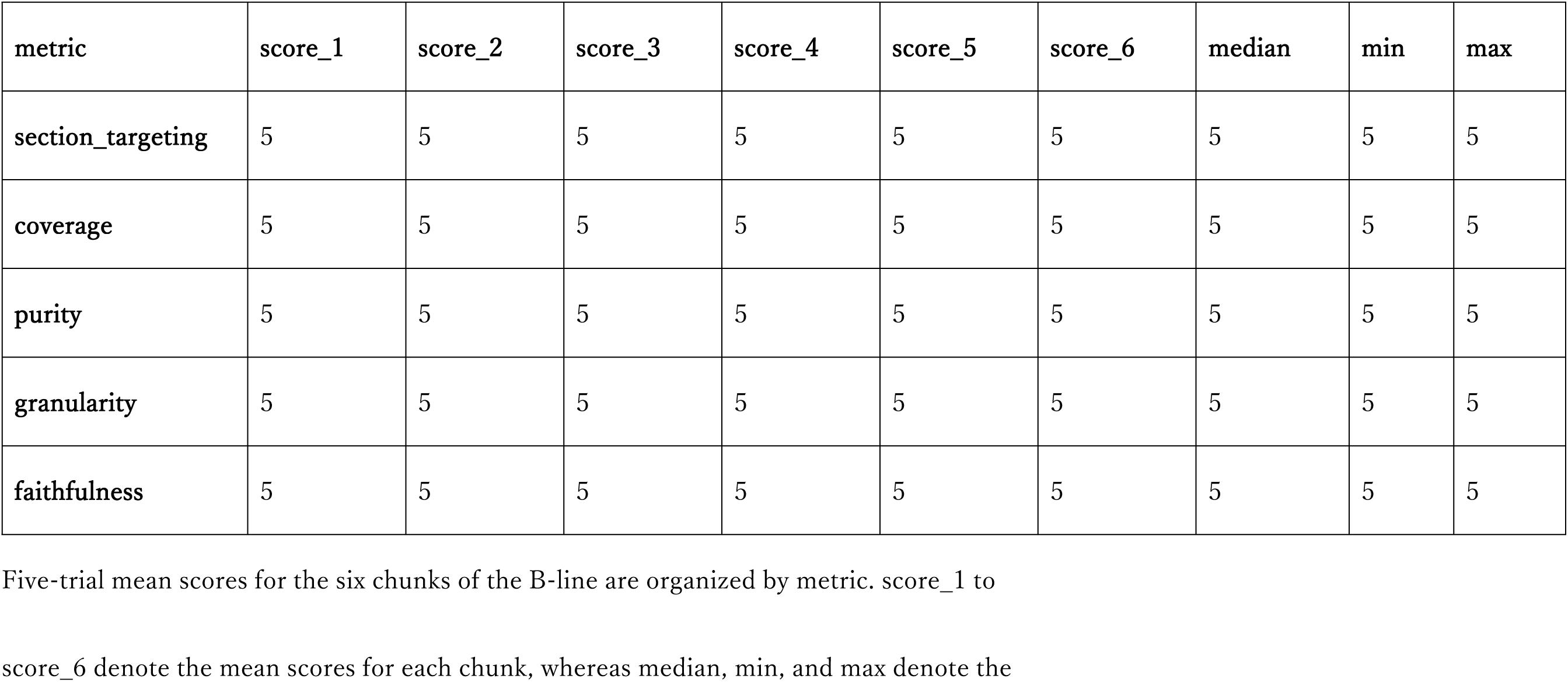
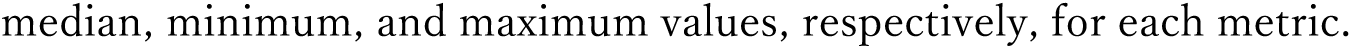
Descriptive statistics of six B-line chunks (Objective section)

The five-trial mean scores of all six B-line chunks recorded a maximum score of 5.0 across all five metrics (Section Targeting, Coverage, Purity, Granularity, and Faithfulness). The median score for each metric was 5.0, with all observations equal to 5.0, indicating no variability among chunks or between trials.

#### 2.7.2 Comparison with A-line Objective

Fig 6 shows six B-line points as a scatter plot overlaid on the box plot of the A-line Objective (n=198) as background.

**Fig 6.**
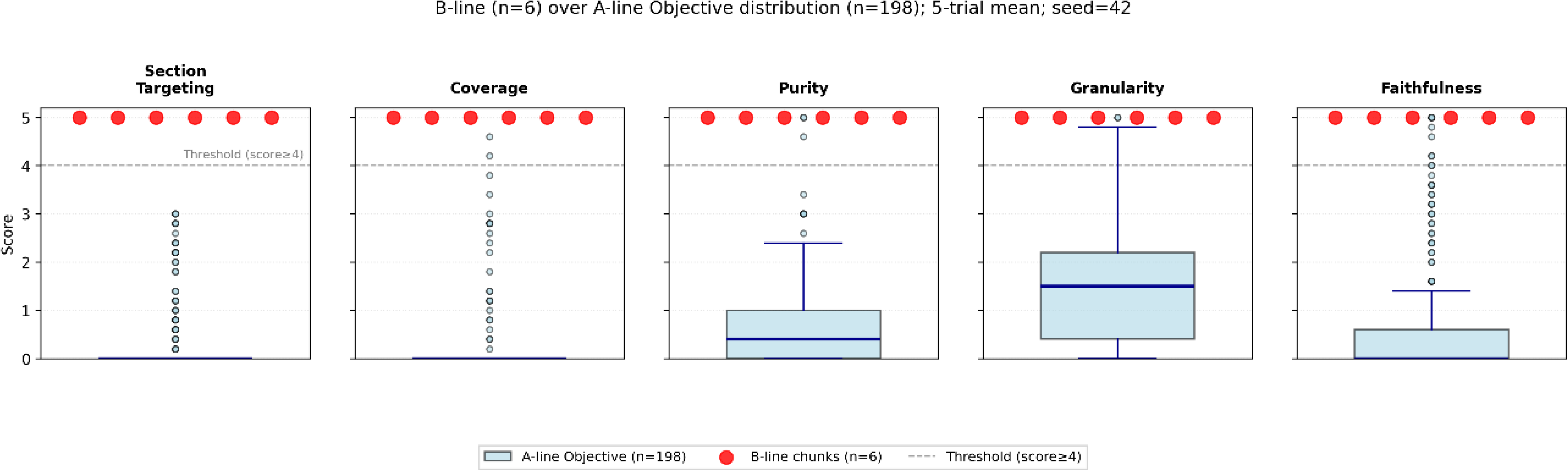
Positional relationship between the A-line Objective distribution and six B-line chunks (five metrics) The blue box plot shows the five-trial mean score distribution of the A-line Objective (n=198). Red dots indicate the five-trial mean scores of the six B-line chunks. The gray dashed line shows the practical utility level threshold (score ≥ 4).

Across all five metrics, the six B-line chunks were positioned above the A-line Objective distribution. For Section Targeting and Coverage, the A-line Objective exhibited a median of 0.0, whereas all six B-line points were positioned at 5.0. For Purity, the A-line Objective had a median of 0.4 and an IQR of 0.0–1.0, whereas all six B-line points exceeded the threshold (score ≥ 4) and were positioned at 5.0. For Granularity and Faithfulness, the six B-line points were located at positions exceeding the upper quartile (Q3) of the A-line Objective distribution.

This result revealed a substantial disparity in sample sizes between the B-line (n=6) and A-line Objective sections (n=198). Because statistical testing was not implemented, no statistical evaluation of between-group differences was performed. The observed pattern is presented as a descriptive finding, and additional verification is required for generalization.

### 2.8 Proof-of-concept: Content appropriateness evaluation (Level 2b)

Descriptive statistics of content appropriateness (Level 2b) were calculated for 99 A-line evaluation chunks (including the query and QE=True condition) for which comparison with GS (SPIRIT 2025 Checklist) was feasible. Violin plots were used to present the descriptive statistics of the three metrics, along with the distribution by section. This analysis was intended as a proof of concept; therefore, statistical testing was not implemented.

#### 2.8.1 GS metrics by section

In the overall descriptive statistics, for the three metrics of overall (n=99), the median of Coverage_gs was 0.0 (IQR: 0.0–1.0), and P(score ≥ 4) was 2.0% (95%CI: 0.6–7.1%). The median of Purity_gs (calculated as 5 − noise) was 3.0 (IQR: 3.0–5.0), and P(score ≥ 4) was 46.5% (95%CI: 37.0–56.2%). Contradiction_gs recorded the maximum score of 5.0 for all chunks, with a median of 5.0 (IQR: 5.0–5.0) and P(score ≥ 4) of 100.0% (95%CI: 96.3–100.0%) (Table 8).

**Table 8.**
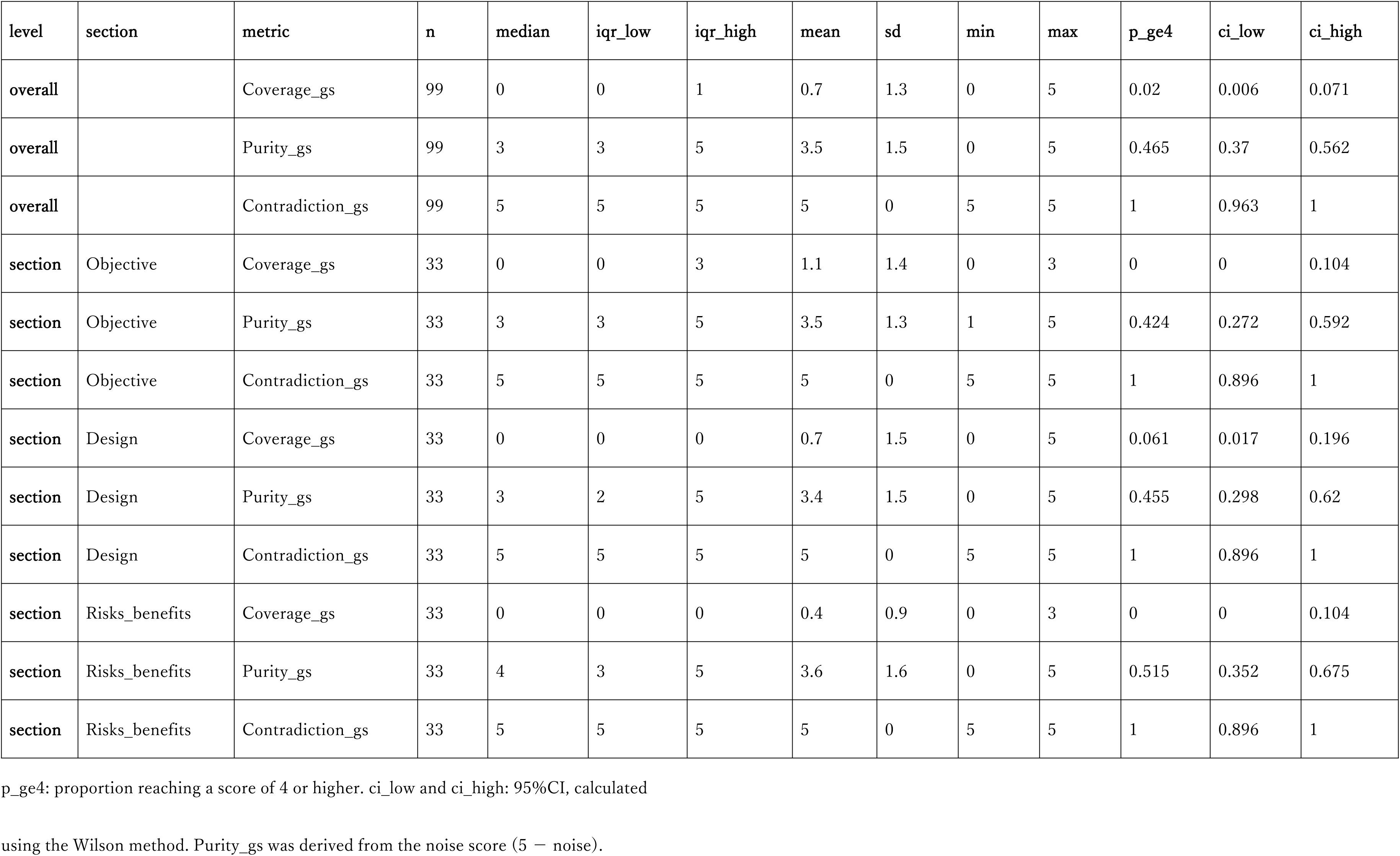
Descriptive statistics by section of Level 2b evaluation (content appropriateness) (n=99)

In the descriptive statistics by section, for each section (each with n=33), the results were as follows: For Coverage_gs, Objective yielded a median of 0.0 (IQR: 0.0–3.0) and P(score ≥ 4) of 0.0% (95%CI: 0.0–10.4%), Design yielded a median of 0.0 (IQR: 0.0–0.0) and P(score ≥ 4) of 6.1% (95%CI: 1.7–19.6%), and Risks_benefits yielded a median of 0.0 (IQR: 0.0–0.0) and P(score ≥ 4) of 0.0% (95%CI: 0.0–10.4%). For Purity_gs, Objective yielded a median of 3.0 (IQR: 3.0–5.0) and P(score ≥ 4) of 42.4% (95%CI: 27.2–59.2%), Design yielded a median of 3.0 (IQR: 2.0–5.0) and P(score ≥ 4) of 45.5% (95%CI: 29.8–62.0%), and Risks_benefits yielded a median of 4.0 (IQR: 3.0–5.0) and P(score ≥ 4) of 51.5% (95%CI: 35.2–67.5%). Contradiction_gs exhibited a median of 5.0 (IQR: 5.0–5.0) and P(score ≥ 4) of 100.0% (95%CI: 89.6–100.0%) for all sections (Table 8).

The distribution of the three metrics across the three sections (Objective, Design, and Risks _benefits) is visualized using violin plots. Coverage_gs was concentrated in the lower part of the distribution across all sections, and chunks exceeding the threshold (score ≥ 4) were rarely observed. Purity_gs exhibited a similar distribution across sections, with a notable thickness of distribution around the median. Contradiction_gs was concentrated at a score of 5.0 for all sections (Fig 7).

**Fig 7.**
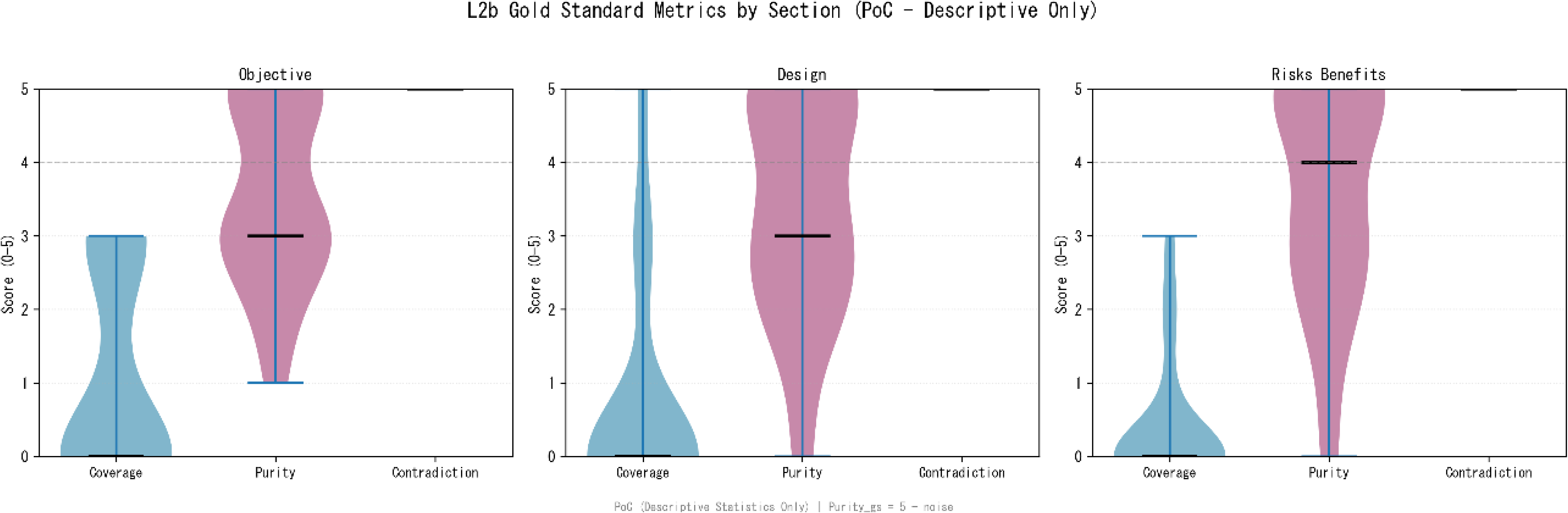
Score distribution by section of L2b Gold Standard metrics (PoC) The score distribution (0–5) of Coverage_gs, Purity_gs, and Contradiction_gs across the three sections of Objective, Design, and Risks_benefits is illustrated using violin plots. The dashed line represents the practical threshold score of 4.0.

#### 2.8.2 Evidence realization rate

Here, we evaluate whether the chunk body text actually includes “evidence text (evidence_trace)” presented by the LLM for each GS element.

For the 99 chunks targeted for the L2b evaluation, recording the evidence_trace for each combination of chunk and GS element resulted in a total of 495 evaluation units. The results of the evidence existence verification (Table 9) indicate that among these units, 422 were entries (evidence_missing) where the LLM response indicated no evidence, such as “Not found,” and did not contain evidence text that could be verified against the body text. The number of entries in which evidence verification was skipped owing to excessive evidence length (evidence_too_long) was 0. For the remaining 73 entries (checkable = 495 − 422 − 0), the presence of the evidence text presented by the LLM within the chunk body text was verified for 59 entries (evidence_found) and denied for 14. The evidence realization rate (evidence_found / checkable) was 59/73 (80.8%).

**Table 9.**
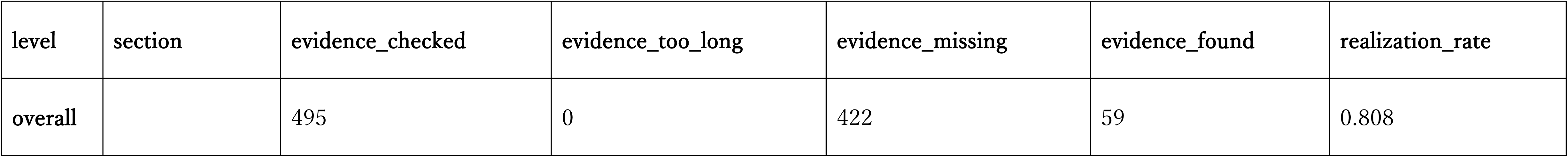
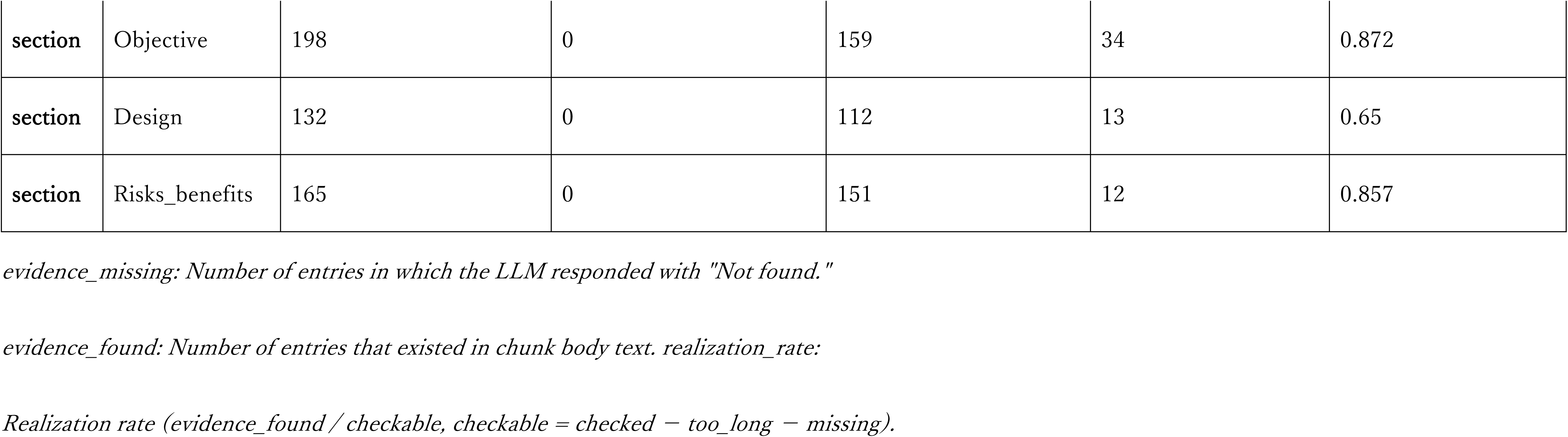
Evidence existence verification results of Level 2b evaluation (n=99 chunks, 495 entries)

The evidence realization rate by section was as follows: Objective: 34/39 (87.2%), Design: 13/20 (65.0%), and Risks_benefits: 12/14 (85.7%).

#### 2.8.3 Chunk characteristics in L2b evaluation

The character count of the 99 chunks targeted for the L2b evaluation was determined. In the descriptive statistics of chunk length, the chunk character count for overall (n=99) had a median of 3,861 characters (IQR: 2,164.5–6,222.0), mean of 4,398.5 characters (SD: 2,988.3), and range of 62–10,343 characters (Table 10).

**Table 10.**
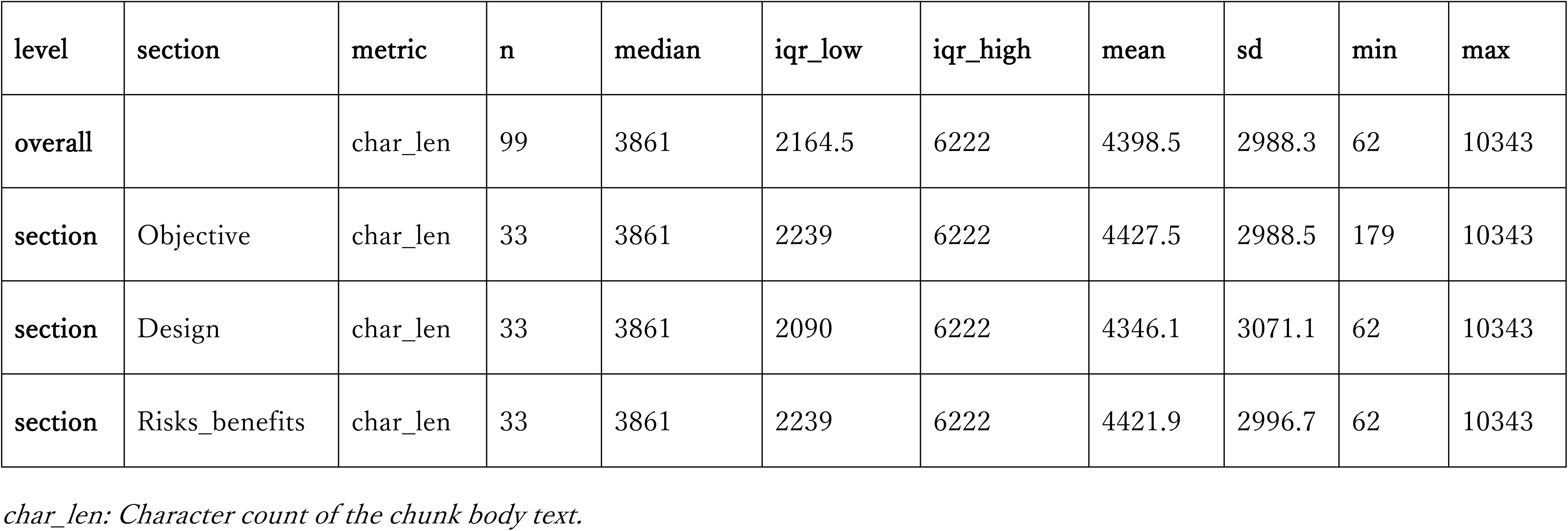
Character count characteristics of chunks targeted for Level 2b evaluation (n=99)

The analysis of chunk length by section revealed median values of 3,861 characters (IQR: 2,239.0–6,222.0) for Objective, 3,861 characters (IQR: 2,090.0–6,222.0) for Design, and 3,861 characters (IQR: 2,239.0–6,222.0) for Risks_benefits. A similar distribution was observed across the different sections (Table 10).

## 3. Discussion

This foundational study quantitatively evaluated the impact of upstream quality (knowledge document quality) in RAG systems on the quality of search results. To our knowledge, few studies have quantified upstream quality at the chunk level, targeting clinical research-related documents in Japanese in the medical field. In the Level 2a evaluation of 594 A-line chunks, the median score across all five metrics was ≤ 2.0, and the proportion of chunks that reached the practical utility level (score ≥ 4) remained below 20%. This result shows that although automatic ingestion functions enable document input without preprocessing, clear limitations arise in the quality of chunks obtained at the retrieval stage if the quality of the upstream document structure and content is insufficient. This finding presents quantitative evidence that challenges the prevalent assumption in practice that “if documents are ingested into a RAG system, it will function sufficiently.” Research aimed at improving RAG performance has thus far focused on optimizing downstream parameters, such as chunk size, search algorithms, and prompt design [6–9]; however, the results of this study suggest that the quality of upstream knowledge can be an important constraint on overall system performance.

The Top-K monotonicity analysis, which verified the relationship between the retrieval rank and chunk quality, revealed statistically significant monotonic trends across all five metrics. The analysis confirmed that the search ranking algorithm tends to place high-quality chunks in the upper positions. However, the effect size (Kendall’s *τ*) did not exceed the Moderate level. Moreover, even in the Rank 1 group, the practical utility level attainment rate was less than half for four metrics, excluding Faithfulness. This result shows that the design expectation that “obtaining top search results is sufficient” is difficult to establish, given the current state of knowledge quality. Although the existence of a statistically significant monotonic trend supports the validity of the search algorithm, it also implies that when the quality of input knowledge is low, the upper limit of output quality is constrained, even at Top-1.

Compared with the chunk size setting of Vertex AI Search (500 tokens), the character count of actually retrieved chunks significantly exceeded the set value at the median. The heading inheritance function (includeAncestorHeadings = true) is intended to add contextual information to chunks, thereby improving the search accuracy [22]. However, the excess character count observed in this study suggests that inherited heading information constitutes a relatively large proportion of the entire chunk, thereby diluting the original information content and consequently decreasing quality scores.

In Level 1 evaluation (independent judgment by three domain experts), the inter-rater agreement was low at *κ* = 0.269, and particularly for the Noise axis, the 95%CI included 0 (*κ* = 0.044, 95%CI: −0.200–0.320). Among the 160 evaluation units, at least one rater assigned an NG judgment in 94 cases (58.8%). However, among these, the judgments were divided among raters in 78 cases (48.8%) (NG×1 or NG×2). This result does not indicate a deficiency in rater capability; rather, it highlights the necessity for refining the evaluation framework designed in this study. This low level of agreement cannot be attributed to a single factor; rather, it reflects the interaction of the quality of knowledge document description, ambiguity of evaluation criteria, and unstructured tacit knowledge that raters individually possess. In other words, this low level of agreement constitutes quantitative evidence that current clinical research manuals are described with sufficient ambiguity to result in varied interpretations among the raters. Therefore, it should be interpreted as a “discovery” of quality issues in the target documents, rather than a “failure” of the evaluation framework.

An analysis of each axis revealed that the judgments were divided among raters in 48.7% of the evaluation units for the Structure axis (n=76), 60.0% for the Granularity axis (n=60), and 87.5% for the Noise axis (n=24). Although the Structure axis showed a relatively high level of agreement (*κ* = 0.306), opinions were approximately evenly divided. By contrast, for the Granularity and Noise axes, the judgments did not agree in the majority of cases. This difference among the axes may reflect the nature of the evaluation items. Items on the Structure axis (including document structure, section division, and version management) are mostly related to the formal features of documents and can be judged relatively objectively. Conversely, the items on the Granularity axis (including level of information detail and sufficiency of definitions) and Noise axis (redundancy and unnecessary information) easily include subjective elements in judgments, such as “what degree is sufficient” and “what is unnecessary.” The quantification of the evaluation criteria and presentation of concrete examples may have been insufficient.

In the B-line proof of concept, structured knowledge (n = 6) achieved the maximum score across all five metrics and was positioned above the A-line Objective distribution. Owing to sample size constraints, statistical testing was not implemented, and causal relationships could not be established based on this result. However, as a directional suggestion that “changing the structure of knowledge greatly changes scores,” this finding provides useful information for future research design.

In the Level 2b evaluation, the practical utility level attainment rate of Coverage_gs was extremely low at 2.0%, whereas that of Contradiction_gs was 100%. This contrasting result suggests that although current RAG systems can retrieve “information that does not contradict the GS,” their ability to comprehensively retrieve “specific instructions required by the GS” is limited. The evidence realization rate (80.8%) shows that much of the evidence text generated by the LLM actually existed in the chunk body text, indicating that the “fabrication” (hallucination) by the LLM was limited. However, among the 495 entries, the LLM responded “Not found” for 422 cases, revealing many areas on the knowledge side where “content that should be written does not exist in the first place.” In other words, the essence of the problem is not likely to be LLM hallucination; rather, content deficiency on the knowledge side is the main constraint factor.

This study highlights the need for further development and dissemination of standardized guidelines for drafting clinical research protocols in Japan. Prior research on IRB review processes [13–15] and materials from the Ministry of Health, Labour and Welfare deliberative councils [16–21] have repeatedly highlighted notable variation in the level of detail, composition, and evaluation perspectives of review comments among CRBs. The variation in manual quality observed in this study may reflect structural issues that share a common origin with the non-uniformity of review processes. Currently, each medical institution maintains its own preparation manuals referring to international standards (such as ICH-GCP) and domestic regulations (including GCP and Clinical Research Act); however, these manuals exhibit variation in their content and composition across organizations. This variation poses challenges in ensuring consistent output quality, even in IRB pre-screening support using RAG systems. The GS (based on the SPIRIT 2025 Checklist) [23] used in the Level 2b evaluation was specifically devised for the research objectives of this study and did not constitute a publicly standardized guideline.

Strengthening the structure of knowledge, formalizing tacit knowledge of experts, and organizing unstructured data to develop a knowledge base infrastructure usable as RAG knowledge, alongside the establishment of standardized guidelines, may improve system accuracy and consistency of the entire review process.

The importance of chunk quality and knowledge base design in RAG has also been highlighted in both general RAG research and surveys of medical RAG [7–9,24]. These studies suggest that performance improvement solely through the optimization of models and prompts is limited and that upstream factors, such as document segmentation and knowledge base development, are important elements for the reliability and safety of RAG. However, within the medical field, particularly in the context of IRB pre-screening for clinical research, reports that quantitatively evaluate the upstream quality of Japanese manuals actually used at the chunk level are limited. To address this gap, this study strategically selected Japanese documents with linguistic complexity, such as the omission of subjects and objects, context-dependent meaning construction through particles, and a mixture of kanji, hiragana, and katakana, and constructed a framework for upstream quality management.

The framework of this study combines a three-tier evaluation framework (Level 1: knowledge quality evaluation, Level 2a: chunk structural quality evaluation, and Level 2b: chunk content appropriateness evaluation), structured knowledge (B-line), and GS development. For implementation, upstream quality management can be incorporated into the IRB pre-screening process by undertaking the following steps: (1) evaluation of knowledge quality (Level 1) and chunk quality (Level 2a, Level 2b); (2) trial of B-line-type structuring; and (3) re-evaluation of content deficiencies through Evidence QA.

This research is intended as a foundational study for designing role allocation between LLM-as-a-Judge and human judges in RAG evaluation. In Level 2a (chunk quality evaluation), the procedure and reliability of LLM-as-a-Judge were verified in Phase 1, given the practical difficulty of evaluating all 600 chunks using human judges. The principal investigator’s visual inspection of all chunks qualitatively ensured the appropriateness of the direction of the LLM judge. These findings suggest the feasibility of a hybrid evaluation framework that combines LLM-as-a-Judge and human judgment, balancing evaluation efficiency and reliability in the medical AI domain.

### 3.1 Limitations

This study had several limitations. First, the evaluation target was limited to the single platform of Google Cloud Vertex AI Search, and generalization to other RAG systems (including Amazon Kendra and Azure AI Search) has not been verified. However, the evaluation framework and measurement procedures established in this study are portable as a platform-independent methodology. Second, the target documents were limited to Japanese study protocol development manuals, and their applicability to multilingual environments and other domains remains unverified. Third, the sample size of the B-line proof of concept (n = 6) was limited, and the target was limited to the Objective section only. Additionally, the sample size of the Level 2b evaluation (n = 99) was limited.

Statistical testing was not implemented. Therefore, the results should be interpreted as descriptive findings, and additional verification is required for generalization. Fourth, compared to the chunk size setting of Vertex AI Search (500 tokens), the character count of actually retrieved chunks significantly exceeded the set value (median of 3,861 characters, range of 62–10,343 characters). The precise technical mechanism responsible for this discrepancy could not be completely identified owing to the lack of publicly available information regarding the logic of accurate token counting, specific algorithm of heading inheritance (includeAncestorHeadings=true), and detailed specifications of tokenization in Japanese text. This technical limitation, attributed to the Vertex AI Search API functioning as a black box, is a common issue in research utilizing cloud-based RAG platforms.

Fifth, the inter-rater agreement for the Level 1 evaluation was *κ* = 0.269, indicating that refining the evaluation framework is a challenge for future research.

Sixth, the results of LLM-as-a-Judge in the Level 2a evaluation were not systematically compared with human judgments rendered by multiple experts in this phase. However, the principal investigator (possessing approximately 18 years of experience in the clinical research support and data management domain and currently engaged in IRB review work) visually inspected all 600 chunks, confirming the absence of qualitative differences from the evaluation results of LLM-as-a-Judge regarding the presence of long chunks and mixing of unrelated sections. Verification based on a quantitative comparison between LLM-as-a-Judge and human judges was identified as a challenge in Phase 2.

Seventh, the GS (based on SPIRIT 2025 Checklist) used in this study was defined for the purposes of this research and does not constitute a publicly standardized guideline.

Additionally, this GS evaluates the comprehensiveness of the structural elements of study protocols rather than directly evaluating the scientific and ethical validity of the protocol content. The establishment of evaluation criteria for validity assessment was identified as a challenge in Phase 2 and beyond.

Eighth, the evidence realization rate of 80.8% was derived from 73 cases, excluding 422 cases where the LLM responded “Not found” among 495 entries, leaving 73 cases as the denominator. This metric differs from the ratio to all entries (59/495 = 11.9%).

Ninth, this study, as a Phase 1 foundational study, focused on grasping measurement properties and did not verify the causal relationship between chunk quality and overall RAG performance (the quality of the final generated output).

Tenth, in this study, LLM-as-a-Judge was implemented using a single model (GPT-4o), and no comparison among models was performed.

### 3.2 Future directions

Based on the findings of this study, the following research directions are proposed as future challenges.

The following four stages will be implemented as a stepwise approach in Phase 2. In the first stage, the 78 items, in which evaluations were divided, will be analyzed by axis, and through the formation of expert consensus using the Delphi consensus method [25], quantitative criteria and judgment indicators will be established for items requiring subjective judgment. In the second stage, structured evaluation manuals, including typical examples and boundary cases, will be developed, and rater training programs will be introduced.

Incorporating these evaluation manuals as a common anchor in rater training and LLM judge prompts will aim to improve the consistency of both human and LLM judges. In the third stage, expert evaluation will be re-implemented with improved evaluation criteria, and improvement of inter-rater agreement (target: *κ* ≥ 0.60, Substantial) will be verified. In the fourth stage, LLM-as-a-Judge will be introduced for knowledge quality evaluation of items where people easily become confused (particularly the Granularity and Noise axes) [26], and the evaluation results of humans and LLMs will be quantitatively compared.

Furthermore, implementing LLM-as-a-Judge using multiple LLM models and verifying the differences in evaluation characteristics among the models will aim to construct a more robust evaluation system. Ultimately, the objective will be to construct an evaluation framework in which human experts and LLMs cooperate.

To verify knowledge and chunk quality, the target of the B-line will be broadened beyond the Objective section to include the Design and Risks_benefits sections, and hypothesis verification research will be conducted using a larger-scale sample. Based on these results, a plan will be established to progressively advance the structural strengthening that targets the full text. Regarding the Level 2a evaluation, Phase 2 will introduce both LLM-as-a-Judge and multiple human experts, and their evaluation results will be quantitatively compared to clarify the validity of LLM-as-a-Judge as a measurement instrument and the appropriate role allocation in practical implementation. Additionally, a comparative experiment based on the presence or absence of heading inheritance settings is planned to verify the causal relationship between chunk length excess and quality decrease.

From a long-term perspective, in pursuit of establishing standardized guidelines for study protocol preparation in Japan, we plan to verify the extent to which advancing the formalization of tacit knowledge that each expert possesses, structuring unstructured documents, and developing a knowledge base infrastructure usable as RAG knowledge contribute to improving system accuracy. Through these efforts, we aim to construct a knowledge base to facilitate the implementation of validity-assessment RAG.

### 3.3 Conclusion

This study quantitatively evaluated the importance of upstream quality (knowledge document quality) in RAG systems. Given the current knowledge quality, even for top-ranked chunks in search, the proportion reaching the practical utility level is limited. This suggests that performance improvement through parameter optimization alone is limited. In this Phase 1 foundational study, we successfully designed a three-tier evaluation framework, confirmed the reliability of LLM-as-a-Judge, suggested directions for improvement through structured knowledge, and established a foundation for hybrid evaluation design involving both LLM-as-a-Judge and human judges. These findings highlight the importance of upstream quality management that precedes downstream optimization in the implementation of RAG systems utilizing medical AI. The three-tier evaluation framework established in this study and the proof of concept of structured knowledge can function as a foundation for the formalization of tacit knowledge of review committee members and the development of validity-assessment RAG for protocol content in the future.

## 4. Materials and methods

### 4.1 Ethics statement

This study targeted only publicly available documents for analysis and did not include human subjects or patient data. The expert evaluation in Level 1 described later was performed by the authors and collaborating researchers as part of system verification and did not correspond to medical research targeting humans; therefore, approval by the IRB was not required.

### 4.2 Study design and overview

As a foundational study (Phase 1) of the RAG process, this research aimed to measure the structural impact of the knowledge base quality on the retrieval result quality. As the technical scope, the search function (Search) of Google Cloud Vertex AI Search was targeted, and only the results when SearchResultMode=CHUNKS was specified in the search API request were analyzed. This mode extracted chunks (original text fragments) from the data store and returned them as they were, without engaging in summarization or answer generation (Generation) processing through LLMs [22,27]. This approach intentionally excluded variation factors from the subsequent generation phase, and the evaluation focused on the impact of knowledge quality on retrieval results (chunks).

The quality evaluation in this study was conducted at three complementary levels to comprehensively grasp the upstream quality of RAG:

1. Level 1 (Knowledge quality evaluation): Three experts independently judged the document quality for each site on three axes and 40 items.
2. Level 2a (Structural quality evaluation): LLM-as-a-Judge evaluated the structural quality of the retrieved chunks based on five metrics.
3. Level 2b (Content appropriateness evaluation): A GS for research use was created, and proof of concept for content quality evaluation was performed.

In this study (Phase 1), the measurement properties (test–retest reliability, inter-rater agreement, and score distribution) of these evaluation frameworks were quantitatively grasped to clarify the requirements for metrics and procedures required for upstream quality management in RAG. The refinement of content validity and the association with overall RAG performance are designated as verification tasks in Phase 2.

### 4.3 Data sources, ingestion, and retrieval

#### 4.3.1 Data sources

This study targeted Japanese clinical research-related documents. Japanese has linguistic characteristics, such as (1) frequent omission of subjects and objects and high context dependence, (2) a tendency for ambiguity in meaning interpretation due to polysemy of particles, and (3) frequent variation in the orthography of medical terms owing to the use of a mixture of kanji, hiragana, and katakana. These characteristics complicate the interpretation process in document analysis, chunking, and meaning extraction by LLMs. Therefore, they are suitable as targets for evaluating the impact of upstream quality in RAG systems.

Regarding knowledge documents, Japanese clinical research protocol development manuals that were published on official websites and freely downloadable as of June 24, 2025 were targeted. Based on selection criteria (Japanese description, structured documents of 30 pages or more, inclusion of specific sections, and non-checklist format), four manuals were selected, and the file names were anonymized, ranging from site-01 to site-04. Owing to copyright (redistribution rights) restrictions and anonymity protection, this group of documents and the acquisition URLs are not disclosed in this paper. To quantify current knowledge quality (baseline measurement) and further explore the potential for improvement through structuring (proof-of-concept), the following two types of dataset lines, each serving a different role, were constructed:

- A-line (baseline knowledge): a dataset in which the selected four manuals were used as they are without changing the structure of the original text, except for the anonymization of site-identifying information.
- B-line (optimized knowledge): an optimized template newly created in this study and targeting only the “Objective” section, referring to the A-line original text group. To achieve both searchability at retrieval time and semantic interpretability by LLMs, structuring was performed in key-value format. Specifically, for each section, (1) mandatory elements (must_include population, intervention, comparison, primary outcome, secondary outcome, and study phase), (2) documentation guidelines (guidelines), (3) example descriptions (examples), and (4) frequently asked questions and answers were arranged hierarchically, and the design was intended to provide explicit instruction sentences for each GS element.

#### 4.3.2 Ingestion: Data store configuration

Google Cloud Vertex AI Search was adopted for the RAG system infrastructure used in this study. UNSTRUCTURED (unstructured data) was selected as the data store type, and the A-line (four documents) and B-line (one document) were imported as independent data stores. In accordance with the specifications outlined in the official documentation of Google Cloud [22,27–30], the ingestion process in Vertex AI Search involves a series of processes in which documents are chunked (divided) and stored as a search index, including vector embeddings. In this study, to strictly evaluate the impact of the independent variable (knowledge quality), the following technical factors within the ingestion settings were fixed:

- Unification of file format: To ensure reproducibility of chunking processing and stability of layout analysis, all knowledge documents were unified to PDF format. As of the technical selection reference date for this study (September 10, 2025), the layout analysis function of Vertex AI Search was generally available (GA, Generally Available) for PDF format, whereas support for Word format (.docx) remained at the Preview stage [29, 30]. To mitigate any potential bias introduced by the instability of Preview functions on experimental results, the GA-completed PDF format was adopted.
- Fixation of chunk generation settings: The layout-based chunking

(LayoutBasedChunkingConfig) function was used, and the main parameters were fixed as follows:

- chunkSize: 500 tokens
- includeAncestorHeadings: true (enabling inheritance of context of ancestor headings)
- defaultParsingConfig: layoutParsingConfig (using layout parser) [22,28]

Because the documents were machine-readable PDFs including text layers, the use of an OCR parser was unnecessary.

The process of ingestion into the data store was automated using the official Python client library (API) to ensure reproducibility and reduce human operational errors.

#### 4.3.3 Query design for retrieval

Queries were designed to retrieve and extract chunk groups that become targets of L2a and L2b evaluations. Three target sections (Objective, Design, Risks_benefits) were selected, and three query types (Write: enquiring about the documentation methods, Include: enquiring about the content to include, and Examples: enquiring about example descriptions) were created. For the A-line, nine queries consisting of combinations of “three sections × three query types” were designed, targeting four sites (site-01 to site-04), and for the B-line, three queries consisting of combinations of “one section (Objective) × three query types” were designed targeting one knowledge source. Details of the queries are included in S1 Text.

#### 4.3.4 Retrieval: Chunk retrieval settings

To execute the designed queries and retrieve and extract (Retrieval) evaluation target chunks, the search method of Google Cloud Vertex AI Search API was used, and processing was automated using a dedicated Python script. The parameters of the search API request were set based on the official reference of Google Cloud [27] and the experimental conditions of this study.

The parameters other than Query Expansion (QE) were fixed, and only QE was switched and executed under two conditions: AUTO (enabled) and DISABLED (disabled).

The settings of each parameter are as follows:

- contentSearchSpec.searchResultMode = CHUNKS: Without performing answer generation using generative AI (answer API or summarization function), chunks of the knowledge source were designated as retrieval targets.
- contentSearchSpec.chunkSpec.numPreviousChunks = 0 and numNextChunks = 0: To evaluate each chunk as an independent unit, the automatic concatenation function of preceding and following context was disabled.
- pageSize = 10: As retrieval results, chunks of the top maximum 10 cases (up to 10) of relevance scores were extracted. In the specifications of Vertex AI Search API, pageSize specifies the upper limit value of returned results, and when the total number of chunks of target documents does not reach the set value, the actual number of retrievals becomes less than 10 cases [27].
- queryExpansionSpec.condition = {AUTO, DISABLED}: Regarding the query expansion function, retrieval was executed under two conditions: enabled (AUTO) and disabled (DISABLED).
- spellCorrectionSpec.mode = AUTO: The spell correction function of queries used the standard setting (automatic).

For the A-line, 18 patterns of “nine queries × two QE conditions” were executed, whereas for the B-line, six patterns of “three queries × two QE conditions” were executed.

### 4.4 Knowledge quality evaluation framework (Human evaluation)

#### 4.4.1 Framework design

In this study, the quality of knowledge documents ingested into Vertex AI Search was defined as “LLM readability,” which indicates whether LLMs can analyze, chunk (divide), and use for retrieval the document content without misunderstanding. To systematically grasp this characteristic, a knowledge quality evaluation framework comprising three axes—Structure, Granularity, and Noise—was designed.

#### 4.4.2 Theoretical rationale for design

The design of the framework was derived deductively based on the official technical specifications of Vertex AI Search and prior findings of information retrieval and RAG research.

- Structure axis: In addition to reports indicating that chunking that preserves the chapter–section structure and layout enhances RAG performance, the utilization of layoutBasedChunkingConfig and SearchResultMode = CHUNKS is recommended in the specifications of Vertex AI Search [6–8,22,24,27,28,31].
- Granularity axis: In this framework, the Granularity axis was defined not as the physical size of information (chunk size) but as the semantic atomicity (detail, clarity, and judgeability of information). Specifically, it refers to the level at which LLMs can grasp structural elements without misunderstanding the meaning, such as abbreviation definitions, explicit statement of mandatory information, concreteness, and clarity of Japanese expressions. This definition is consistent with recent RAG research indicating that granularity of information affects faithfulness and usability of generated outputs [6–8,24].
- Noise axis: Seminal works in the information retrieval field and recent studies have shown that the coexistence of unnecessary comments, unfilled placeholders, and irrelevant URLs, among others, degrades retrieval performance [32–35].

Integrating the aforementioned elements, the model was systematized as three-dimensional (Structure–Granularity–Noise) based on the settings of Vertex AI Search. This framework fixed (i) the official technical specifications of Vertex AI Search; (ii) prior research, including peer-reviewed articles and preprints on information retrieval and RAG; and (iii) evaluation procedures determined in a pre-registered manner before the start of this study. Note that no post-hoc modifications were performed at the discretion of the researchers.

The English translation version of this evaluation framework (definitions of three axes; complete list of 13 evaluation items and 40 Evaluation Criteria; and representative examples of Rationale, Positive/Negative Examples, and Scope used for rater training) is included in Supporting Information (S2 Text).

#### 4.4.3 Evaluation targets and procedures

The knowledge documents of the four A-line sites (site-01 to site-04) were used as evaluation targets. The Structure and Noise axes targeted all documents (all pages) of each site, and the evaluation of the Granularity axis was limited to the “Objective,” “Design,” and “Risks_benefits” sections of the study protocols. The B-line was excluded from this evaluation.

Three experts in clinical research support, biostatistics, and clinical research data management performed independent evaluations. Each rater applied 13 evaluation items (40 Evaluation Criteria) to documents of the four sites and judged using the OK/NG (binary) method (total of 480 judgments: 40 criteria × four sites × three persons).

Because this study is exploratory in nature, no unification through consensus was performed for items where judgments among raters were divided, and independent judgments were recorded as they are.

Prior to evaluation, the three experts received pre-training (calibration) using the Japanese version of the evaluation framework (Rationale, Evaluation Criteria, Positive/Negative Examples, and Scope of 13 evaluation items), and evaluation was performed after unifying interpretation of evaluation criteria based on Examples.

### 4.5 Level 2a: Chunk structural quality evaluation (LLM-as-a-Judge)

#### 4.5.1 Framework and approach of evaluation

In this study, the structural quality of the output of the retrieval component (Retrieval) was evaluated by collating generated chunks with the corresponding original text paragraphs (Reference) in the original document, and the validity of the chunk processing was quantified.

The LLM-as-a-Judge paradigm [26,36], which has been established in recent natural language processing research, was adopted as the evaluation method. This method enables evaluation with a high correlation with human experts by assigning task-specific evaluation rubrics (scoring criteria) to high-performance LLMs such as GPT-4. In this study, rather than using a simple binary judgment, a holistic evaluation (Holistic Evaluation) was performed using a 0–5-point Likert scale, with detailed definitions given to each metric. This design aimed to capture fine differences in chunk quality.

#### 4.5.2 Target chunks for evaluation

Among the retrieved and extracted chunks, the chunks extracted from the A-line and B-line were designated targets of the L2a evaluation.

#### 4.5.3 Evaluation metrics

To comprehensively evaluate chunk quality, the following five metrics were defined, each rated on a scale of 0–5 points. These metrics are adaptations of concepts [24,37–39] proposed in recent research on document processing and RAG evaluation to the context of this study, which focuses on the “quality evaluation of chunks generated by commercial retrieval systems.” Detailed prompts and scoring criteria are included in Supporting Information S3.

##### 1. Section Targeting

The extent to which the main content of chunks accurately belongs to the designated sections (Objective/Design/Risks_benefits) was evaluated based on the concept of “Structural Preservation” advocated in Vision-Guided Chunking by Tripathi et al. [37]. Mixing of the table of contents or inclusion of irrelevant sections was subject to point deduction.

##### 2. Coverage

In alignment with the concept of “Information Completeness” proposed by Tripathi et al. [37], the extent to which mandatory information from the corresponding original text paragraphs is comprehensively captured in the chunks was evaluated. A criterion was established whereby five points were awarded for retaining 80% or more of the information and points were deducted according to the degree of information omission.

##### 3. Purity

The concept of “Chunk Purity”, advocated by Verma [38] in the context of S2 Chunking, was adapted. Verma originally defined it as a metric for measuring the homogeneity of clustering. However, this study reinterpreted it as “absence of semantic noise” and evaluated the degree of mixing of headers and footers, page numbers, and fragments of other sections, among others (inverse score with no noise as five points).

##### 4. Granularity

The concepts of “Boundary Detection” and “Adaptive Granularity” proposed in Meta-Chunking by Zhao et al. [24] were applied. This study evaluated whether chunks were cut without regard to context or if their excessive length resulted in a loss of focus. The optimality of these chunks as semantic units was scored on a scale of 0–5 points.

##### 5. Faithfulness

Within the RAGAS framework, the concept of Faithfulness, as defined by Es et al. [39], is typically used to detect “hallucination of generated answers (Hallucination).” In this study, this concept was extended to the preprocessing stage, serving as a metric to evaluate whether extracted chunk texts contain “alteration, contradiction, or omission of the meaning of the original text” due to PDF parsing errors or character corruption.

#### 4.5.4 Implementation of LLM evaluation

L2a evaluation was performed based on the LLM-as-a-Judge paradigm. GPT-4o (OpenAI, model version: gpt-4o-2024-08-06) was used as the judge and executed with deterministic settings (temperature = 0, seed = 42, max_tokens = 2000, top_p = 1.0, frequency_penalty = 0, and presence_penalty = 0).

The evaluation process was automated using scripts, with each chunk × each trial performed as an independent API call.

Each chunk was independently evaluated five times, and after verifying reliability as a measurement instrument (LLM judge) through ICC(2,1), the mean score from five trials (0–5 points) was used for the main analysis.

### 4.6 Level 2b: GS content evaluation (LLM-as-a-Judge)

#### 4.6.1 Framework of evaluation and comparison target (GS)

Existing study protocol development manuals are independently developed by each organization, referencing both domestic and international regulations and guidelines; therefore, their content and structure lack uniformity. Given this diversity, this study required a comparable evaluation framework; therefore, a GS was established as the criterion for content evaluation. The GS defines, at the element level, the “content that should be described at minimum in study protocols (What)” for the three sections—Objective, Design, and Risks_benefits—and has a different role from the B-line, which is the ideal form of the knowledge structure.

During creation, SPIRIT 2025 Checklist [23], which is an international standard, was used as the reference criterion. Through the consensus of three experts, the elements were confirmed by adapting its mandatory items to the practical customs of Japanese clinical research protocols (description tendencies of four A-line sites). These definitions were implemented into the system as structured data in the YAML format of Japanese clinical research protocols (element ID, label, and keyword group) for each section and were referenced during evaluation execution.

#### 4.6.2 Selection of evaluation targets

Because L2b evaluation must measure the degree of agreement with the “content that should be described (What)” as defined by GS, the targets were limited to the Include query type.

Furthermore, to unify the comparison conditions with GS, QE was set to AUTO/on, and only chunks of include × QE-on retrieved from the A-line were targeted.

#### 4.6.3 Evaluation metrics

For each chunk, the fulfillment status of GS elements and the validity of content were evaluated using the following three metrics (each rated on a scale of 0–5 points). Although the metric names in L2b are similar to those in L2a, the difference is that in L2b, the evaluation targets are the degree of content fulfillment for GS elements and presence or absence of contradiction, rather than “structural agreement with the original manual.” Additionally, drawing on the concept of Coverage/Faithfulness proposed in RAGAS [39], the answer evaluation metrics in RAG are extended to GS-based content evaluation. Detailed prompts and scoring criteria are included in Supporting Information S4.

##### 1. Coverage_gs (degree of fulfillment of explicit GS instructions)

For each element ID defined in the GS YAML of each section, the presence of explicit instruction sentences such as “include ∼,” “describe ∼,” and “specify about ∼” in the chunk was judged. This was not considered fulfilled with mere examples (Example) or implicit mention alone, and scoring was performed on a scale of 0–5 points according to the proportion of elements with “explicit instructions” (e.g., 100% = 5 points and 50–79% = 3 points).

##### 2. Purity_gs (inverse score of non-GS content contamination)

Within the content of the entire chunk, the proportion of topics unrelated to GS elements (including explanations of other sections, operational procedures, and general cautions) was evaluated on a scale of 0–5 points. On the prompt, this proportion was evaluated as Noise (noise amount) on a scale of 0–5 points. In this study, Purity_gs was derived from the Noise score as Purity_gs = 5 − Noise, serving as an inverse score representing the “purity of content related to GS.” While Purity of L2a targets structural noise, such as headers/footers, Purity_gs of L2b targets content noise for GS.

##### 3. Contradiction_gs (degree of contradiction with GS definitions)

Based on the Faithfulness metric defined in RAGAS [39], the extent to which descriptions within chunks contradict the definitions of GS elements was evaluated on a scale of 0–5 points. Points were deducted when descriptions clearly differed from the requirements of GS (e.g., describing as optional items that should be mandatory); the case with no contradiction was assigned 5 points, and the case of complete contradiction was assigned 0 points.

#### 4.6.4 Implementation of LLM evaluation

L2b evaluation was performed based on the LLM-as-a-Judge paradigm [26,36], similarly to L2a evaluation. GPT-4o (OpenAI, model version: gpt-4o-2024-08-06) was used as the judge, and evaluation was performed once per chunk using the same model and evaluation settings as those defined for the L2a evaluation. The evaluation process was automated using scripts and executed as an independent API call for each chunk.

In this phase, L2b serves as a proof of concept. Based on the results of L2a reliability analysis a single trial was conducted, and the distribution of the three metrics—Coverage_gs, Purity_gs, and Contradiction_gs—along with the verdict, was analyzed as descriptive statistics.

### 4.7 Statistical analysis

All statistical analyses were performed using Python 3.10.11 (pandas, NumPy, SciPy, pingouin, statsmodels, Matplotlib). Evaluation by LLM-as-a-Judge was performed using deterministic settings (temperature = 0 and seed = 42), and processing using random numbers (bootstrap, permutation test) used the same seed (42) to ensure reproducibility.

#### 4.7.1 Reliability analysis of LLM-as-a-Judge (Level 2a)

To verify the measurement stability in the L2a evaluation, five independent evaluations (total of 2,970 evaluations) were obtained for 594 A-line chunks, and ICC(2,1) (two-way random-effects, absolute agreement, single measurement) was calculated. The 95%CI for the ICC was estimated using the F distribution, as outlined by Shrout and Fleiss [40]. The ICC was calculated for each of the five metrics and for the five-metric mean score (overall ICC). The interpretation of the ICC followed the guidelines proposed by Koo and Li [41], classifying the ICC as Poor (<0.50), Moderate (0.50–0.75), Good (0.75–0.90), and Excellent (>0.90). Before analysis, data quality criteria were set as follows: (1) The ICC of all five metrics is 0.75 or above, (2) each chunk retains all five trials. Only the chunks that met these criteria were used for subsequent analyses. In subsequent analyses, the mean score of five trials was used.

#### 4.7.2 Descriptive statistics for L2a chunk quality

Descriptive statistics (median, IQR, mean, SD, and range) of five metrics (Section Targeting, Coverage, Purity, Granularity, and Faithfulness) were calculated for the 594 A-line chunks that met the ICC criteria.

P(score ≥ 4) was calculated as a metric of practical utility, and the 95%CI was estimated using the Wilson method (without continuity correction) [42].

Stratified descriptive statistics by Section, Site, and Query Expansion (on/off) were calculated. However, no statistical testing of between-group comparisons was performed.

To confirm the distribution of the five metrics, box plots and violin plots by section were created.

#### 4.7.3 Chunk length distribution analysis

The character count distribution of the 594 A-line chunks was evaluated. To quantify the impact of setting parameters (chunkSize = 500 tokens and includeAncestorHeadings = true), descriptive statistics were calculated and visualization was performed.

For each QE (on/off) × section (Objective, Design, Risks_benefits), n, median, IQR, mean, SD, and range were calculated.

The overall distribution was visualized using a histogram, and stratified distributions were visualized using box plots.

#### 4.7.4 Top-K monotonicity analysis (Jonckheere–Terpstra test and Kendall’s *τ*)

To verify the monotonic relationship between retrieval rank (Rank) and chunk quality scores, Rank 1–5 were classified into three groups: Rank 1, Rank 2–3, and Rank 4–5, and Jonckheere–Terpstra test was performed [43, 44]. The p value was calculated using a site-block permutation test (10,000 iterations and seed = 42), which shuffles group labels only within sites. Kendall’s *τ* was calculated as a measure of the effect size, and the bootstrap 95% CI (1,000 iterations) was estimated. To unify the analysis direction, the Rank values were reversed, establishing *τ* > 0 as the hypothesized direction (low Rank → high quality). As supplementary analysis, the overall *τ* of Rank 1–10 was also calculated. For each Rank group, descriptive statistics and P(score ≥ 4) were calculated, and the distribution was visualized using box plots and strip plots.

#### 4.7.5 Level 1: Knowledge quality evaluation (Expert evaluation)

Fleiss’ *κ* was calculated for a total 480 judgments across 160 evaluation points (four sites × 40 items) by three experts, and evaluation was also performed for each axis (Structure, Granularity, and Noise). The 95%CI was estimated using a site-stratified bootstrap (1,000 iterations).

The criteria of Landis and Koch [45] were used for interpretation of Fleiss’ *κ*.

In addition, for each evaluation point, the number of NG judgments (0–3) was tallied, and the distribution was calculated both overall and for each axis. The pattern of variation in the evaluation results was then visualized.

#### 4.7.6 B-line proof-of-concept analysis

The five-trial mean score was calculated for the six B-line chunks of the Objective section and was then descriptively compared by juxtaposition with the distribution of the A-line Objective (n = 198).

For the B-line, the individual scores, median, and range of each metric were calculated, and no statistical testing was performed.

For visualization, a figure was generated in which the score scatter plot of the B-line was overlaid on the box plot of the A-line.

#### 4.7.7 Level 2b statistical analysis

Descriptive statistics for the evaluation results of L2b (Coverage_gs, Purity_gs, Contradiction_gs) were calculated both overall and by section (median, IQR, mean, SD, range, P(score ≥ 4), Wilson 95% CI).

The existence of Evidence was verified through string matching, and the evidence realization rate was calculated.

Descriptive statistics were similarly calculated for the character count (char_len) of each chunk.

Violin plots of section × metric were created to confirm the distribution of the three metrics.

## Data Availability

Data Availability: All data underlying the findings of this study are included in the manuscript and its Supporting Information files. The Supporting Information files and figure source data will be made publicly available via a permanent repository (e.g., Figshare) prior to publication. The original source documents analyzed in this study are publicly available Japanese clinical research protocol development manuals however, the full texts and source URLs are not redistributed due to copyright and anonymization considerations. No personal or identifying information is included.

## Acknowledgments

We thank ChatGPT (OpenAI) and Claude (Anthropic) for assistance with English translation and editing of Japanese-language research materials, including manuscript drafts and example texts. These tools were also used to support the drafting of Python scripts for data processing and analysis. All AI-assisted outputs, including code and translations, were carefully reviewed, tested, and validated by the author prior to use.

We also thank Editage (Cactus Communications) for providing professional English-language editing of the final manuscript.

The author takes full responsibility for the accuracy, integrity, and interpretation of the manuscript content.

## Supporting information captions

S1 Table. Descriptive statistics by retrieval rank.

S1 Fig. Proportion achieving score ≥ 4 across retrieval ranks (Rank 1–10).

S2 Fig. Score distribution by retrieval rank.

S1 Text. Query Catalog for Chunk Retrieval

S2 Text. Knowledge Quality Evaluation Framework

S3 Text. Level 2a Evaluation Prompt and Scoring Criteria (LLM-as-a-Judge; Structural Quality)

S4 Text. Level 2b Evaluation Prompt and Scoring Criteria (LLM-as-a-Judge; Content Appropriateness)

## References

1. Roustan D, Bastardot F. The clinicians’ guide to large language models: a general perspective with a focus on hallucinations. Interact J Med Res. 2025;14:e59823. doi: 10.2196/59823.

2. Lewis P, Perez E, Piktus A, Petroni F, Karpukhin V, Goyal N, et al. Retrieval-augmented generation for knowledge-intensive NLP tasks. Adv Neural Inf Process Syst. 2020;33:9459–9474.

3. Zhu Z, Zhang Y, Zhuang X, Zhang F, Wan Z, Chen Y, et al. Can we trust AI doctors? A Survey of medical hallucination in large language and large vision-language models. Findings of the Association for Computational Linguistics: ACL 2025. 2025; 6748–6769. Available from: https://aclanthology.org/2025.findings-acl.350.pdf

4. Granstedt J, Kc P, Deshpande R, Garcia V, Badano A. Hallucinations in medical devices. Artif Intell Life Sci. 2025;8:100145. doi: 10.1016/j.ailsci.2025.100145.

5. Hiriyanna S, Zhao W. Multi-layered framework for LLM hallucination mitigation in high-stakes applications: A Tutorial. Comput. 2025;14:332. doi: 10.3390/computers14080332.

6. Bhat SR, Rudat M, Spiekermann J, Flores-Herr N. Rethinking chunk size for long-document retrieval: a multi-dataset analysis. arXiv [Preprint]. 2025 [cited 2025 Nov 3]. Available from: https://arxiv.org/abs/2505.21700

7. Wang Z, Gao C, Xiao C, Huang Y, Si S, Luo K, et al. Document segmentation matters for retrieval-augmented generation. Findings of the Association for Computational Linguistics: ACL 2025. 2025; 8063–8075.

8. Liu Z, Simon C-E, Caspani F. Passage segmentation of documents for extractive question answering. arXiv [Preprint]. 2025 [cited 2025 Nov 3]. Available from: https://arxiv.org/abs/2501.09940

9. Oche AJ, Folashade AG, Ghosal T, Biswas A. A systematic review of key retrieval-augmented generation (RAG) systems: progress, gaps, and future directions. arXiv [Preprint]. 2025 [cited 2025 Nov 3]. Available from: https://arxiv.org/abs/2507.18910

10. Jimeno-Yepes A, You Y, Milczek J, Laverde S, Li L. Financial report chunking for effective retrieval-augmented generation. arXiv [Preprint]. 2024 [cited 2025 Nov 3]. Available from: https://arxiv.org/abs/2402.05131

11. Saha B, Saha U, Malik M Z. QuIM-RAG: Advancing retrieval-augmented generation with inverted question matching for enhanced QA performance. IEEE Access. 2024;12:185401–185410. doi: 10.1109/ACCESS.2024.3513155.

12. Ge X, Murtaza S, Cortez A, Alemzadeh H. Expert-guided prompting and retrieval-augmented generation for emergency medical service question answering. arXiv [Preprint]. 2025 Nov [cited 2025 Dec 1]. Available from: https://arxiv.org/html/2511.10900v1

13. Larson E, Bratts T, Zwanziger J, Stone P. A survey of IRB process in 68 U.S. hospitals. J Nurs Scholarsh. 2004;36(3):260–264. doi: 10.1111/j.1547-5069.2004.04047.x.

14. Dyrbye LN, Thomas MR, Mechaber AJ, Eacker A, Harper W, Massie FS Jr, et al. Medical education research and IRB review: an analysis and comparison of the IRB review process at six institutions. Acad Med. 2007;82(7):654–660. doi: 10.1097/ACM.0b013e318065be1e.

15. Abbott L, Grady C. A systematic review of the empirical literature evaluating IRBs: what we know and what we still need to learn. J Empir Res Hum Res Ethics. 2011;6(1):3–19. doi: 10.1525/jer.2011.6.1.3.

16. Ministry of Health, Labour and Welfare (MHLW). Minutes of the 2nd Clinical Research Subcommittee Meeting of the Health Science Council. 2017 Aug 31 [cited 2025 Jun 15]. Available from: https://www.mhlw.go.jp/stf/shingi2/0000179030.html. Japanese.

17. MHLW. Minutes of the 10th Clinical Research Subcommittee Meeting of the Health Science Council. 2019 Feb 15 [cited 2025 Jun 15]. Available from: https://www.mhlw.go.jp/stf/newpage_03963.html. Japanese.

18. MHLW. Minutes of the 34th Clinical Research Subcommittee Meeting of the Health Science Council. 2024 Jan 31 [cited 2025 Jun 15]. Available from: https://www.mhlw.go.jp/stf/newpage_38195.html. Japanese.

19. MHLW. Document 1 from the 34th Clinical Research Subcommittee Meeting of the Health Science Council: On certified clinical research review boards. 2024 Jan 31 [cited 2025 Jun 15]. Available from: https://www.mhlw.go.jp/stf/newpage_37286.html. Japanese.

20. MHLW. Minutes of the 35th Clinical Research Subcommittee Meeting of the Health Science Council. 2024 Aug 8 [cited 2025 Jun 15]. Available from: https://www.mhlw.go.jp/stf/newpage_43555.html. Japanese.

21. MHLW. Documents 1–6 from the 35th Clinical Research Subcommittee Meeting of the Health Science Council: On certified clinical research review boards. 2024 Aug 8 [cited 2025 Jun 15]. Available from: https://www.mhlw.go.jp/stf/newpage_42147.html. Japanese.

22. Google Cloud. Parse and chunk documents | Vertex AI Search. Google Cloud Documentation. 2025 [cited 2025 Dec 1]. Available from: https://docs.cloud.google.com/generative-ai-app-builder/docs/parse-chunk-documents

23. Chan AW, Boutron I, Hopewell S, Moher D, Schulz KF, Collins GS, et al. SPIRIT 2025 statement: updated guideline for protocols of randomized trials. Nat Med. 2025;31(6):1784–1792. doi: 10.1038/s41591-025-03668-w.

24. Zhao J, Ji Z, Feng Y, Qi P, Niu S, Tang B, et al. Meta-chunking: Learning efficient text segmentation via logical perception. arXiv [Preprint]. 2024 Nov 25 [cited 2025 Dec 1]. Available from: https://arxiv.org/abs/2410.12788

25. Hsu CC, Sandford BA. The Delphi technique: Making sense of consensus. Pract Assess Res Eval. 2007;12(10):1–8. doi: 10.7275/pdz9-th90.

26. Zheng L, Chiang WL, Sheng Y, Zhuang S, Wu Z, Zhuang Y, et al. Judging LLM-as-a-Judge with MT-bench and chatbot arena. arXiv [Preprint]. 2023 [cited 2025 Dec 1]. Available from: https://arxiv.org/abs/2306.05685

27. Google Cloud. Vertex AI Search REST API Reference: projects.locations.collections.engines.servingConfigs.search [Internet]. [cited 2025 Sep 10]. Available from: https://cloud.google.com/generative-ai-app-builder/docs/reference/rest/v1/projects.locations.collections.engines.servingConfigs/search

28. Google Cloud. Vertex AI Search REST API Reference: projects.locations.collections.dataStores (LayoutBasedChunkingConfig) [Internet]. [cited 2025 Sep 10]. Available from: https://cloud.google.com/generative-ai-app-builder/docs/reference/rest/v1/projects.locations.collections.dataStores

29. Google Cloud. Prepare data for ingestion | Vertex AI Search [Internet]. [cited 2025 Sep 10]. Available from: https://cloud.google.com/generative-ai-app-builder/docs/prepare-data

30. Google Cloud. Vertex AI Agent Builder Release Notes [Internet]. [cited 2025 Sep 10]. Available from: https://cloud.google.com/generative-ai-app-builder/docs/release-notes

31. Microsoft. Retrieval-augmented generation in Azure AI Search [Internet]. Microsoft Learn. 2025 [cited 2025 Dec 1]. Available from: https://learn.microsoft.com/en-us/azure/search/retrieval-augmented-generation-overview

32. Manning CD, Raghavan P, Schütze H. Introduction to information retrieval. Cambridge: Cambridge University Press; 2008.

33. Kohlschütter C, Fankhauser P, Neumann G. Boilerplate detection using shallow text features. In: Proceedings of the Third ACM International Conference on Web Search and Data Mining (WSDM ’10). New York: ACM; 2010. p. 441–450.

34. Vogels T, Ganea OE, Eickhoff C. Web2Text: Deep structured boilerplate removal. In: Pasi G, Piwowarski B, Azzopardi L, Hanbury A, editors. Advances in information retrieval (ECIR 2018). Lecture Notes in Computer Science, vol. 10772. Cham: Springer; 2018. p. 361–375.

35. Fernández-Pichel M, Prada-Corral M, Losada DE, Pichel JC, Gamallo P. An unsupervised perplexity-based method for boilerplate removal. Nat Lang Eng. 2024;30(1):132–149. doi: 10.1017/S1351324923000049.

36. Liu Y, Iter D, Xu Y, Wang S, Xu R, Zhu C. G-Eval: NLG evaluation using GPT-4 with better human alignment. arXiv [Preprint]. 2023 [cited 2025 Dec 1]. Available from: https://arxiv.org/abs/2303.16634

37. Tripathi V, Odapally T, Das I, Allu U, Ahmed B. Vision-guided chunking is all you need: Enhancing RAG with multimodal document understanding. arXiv [Preprint]. 2025 [cited 2025 Dec 2]. Available from: https://arxiv.org/abs/2506.16035

38. Verma P. S2 chunking: A hybrid framework for document segmentation through integrated spatial and semantic analysis. arXiv [Preprint]. 2025 [cited 2025 Dec 2]. Available from: https://arxiv.org/abs/2501.05485

39. Es S, James J, Espinosa-Anke L, Schockaert S. RAGAS: Automated evaluation of retrieval-augmented generation. arXiv [Preprint]. 2023 [cited 2025 Dec 2]. Available from: https://arxiv.org/abs/2309.15217

40. Shrout PE, Fleiss JL. Intraclass correlations: uses in assessing rater reliability. Psychol Bull. 1979;86(2):420–428. doi: 10.1037/0033-2909.86.2.420.

41. Koo TK, Li MY. A guideline of selecting and reporting intraclass correlation coefficients for reliability research. J Chiropr Med. 2016;15(2):155–163. doi: 10.1016/j.jcm.2016.02.012.

42. Wilson EB. Probable inference, the law of succession, and statistical inference. J Am Stat Assoc. 1927;22(158):209–212. doi: 10.1080/01621459.1927.10502953.

43. Jonckheere AR. A distribution-free k-sample test against ordered alternatives. Biometrika. 1954;41(1–2):133–145. doi: 10.2307/2333011

44. Terpstra TJ. The asymptotic normality and consistency of Kendall’s test for trend when ties are present in one ranking. Indag Math. 1952;14:327–333. doi: 10.1016/S1385-7258(52)50043-X.

45. Landis JR, Koch GG. The measurement of observer agreement for categorical data. Biometrics. 1977;33(1):159–174. doi: 10.2307/2529310.

